# An explainable transformer model learning from entire treatment timelines for pan-cancer risk profiling across healthcare systems

**DOI:** 10.64898/2026.07.24.26358838

**Authors:** Philipp Keyl, Niklas Kiermeyer, Jonah Bosserhoff, Tim Lenfers, Thibault Niederhauser, Bowen Fan, Thomas Schnake, Simon Schallenberg, Fabio Aubele, Solveig Kuss, Mina Jamshidi Idaji, Philipp Jurmeister, Moon Kim, Sebastian Bauer, Nikolaos Bechrakis, Michael Forsting, Dagmar Führer-Sakel, Martin Glas, Viktor Grünwald, Boris Hadaschik, Ken Herrmann, Stefan Kasper, Rainer Kimmig, Stephan Lang, Ina Pretzell, Tienush Rassaf, Alexander Roesch, Jens T. Siveke, Maja Guberina, Ulrich Sure, Marc Wichert, Michael Ingrisch, Kristian Unger, Jürgen Behr, Daniel Teupser, Christian G. Stief, Julia Mayerle, Nadia Harbeck, Amanda Tufman, Jens Ricke, Lars H. Lindner, Siegfried Priglinger, Günter Höglinger, Sven Mahner, Martin Canis, Lucie Heinzerling, Christine Spitzweg, Alpaslan Tasdogan, Matthias Totzeck, Anja Welt, Marcel Wiesweg, C. Benedikt Westphalen, Reinhard Thasler, Fady Albashiti, Grégoire Montavon, Nicola Miglino, Zsolt Balázs, Michael von Bergwelt-Baildon, Volker Heinemann, Claus Belka, Sylvia Hartmann, Andreas Wicki, Felix Nensa, Dirk Schadendorf, Michael Krauthammer, Klaus-Robert Müller, Martin Schuler, Frederick Klauschen, Jens Kleesiek, Julius Keyl

**Author notes:** These authors contributed equally (Philipp Keyl, Niklas Kiermeyer). These authors jointly supervised this work (Frederick Klauschen, Jens Kleesiek, Julius Keyl).

## Abstract

Cancer outcomes vary widely between individual patients, each accumulating an irregular record of treatments, diagnoses, measurements, and complications. Current prognostic models reduce this complexity into a single snapshot, focus on narrow clinical settings, and rarely generalize across hospitals. Here we introduce **Chronicle**, an explainable transformer that learns from entire patient trajectories to predict diverse clinical outcomes throughout the disease course while capturing both short- and long-term temporal dependencies.

Trained on 53.7 million longitudinal data points from 51,711 patients spanning 67 cancer types, Chronicle operates natively on irregular data without imputation and jointly predicts eight endpoints within a flexible framework adaptable to additional outcomes. Chronicle outperformed cross-sectional models for overall survival prediction (C-index 0.84 vs 0.76-0.79), stratified patients more accurately than established prognostic systems, including TNM stage, and predicted seven adverse event and transfusion endpoints (AUC 0.80-0.92). Applied without retraining to 69,341 patients in Germany, Switzerland, and the United States, Chronicle generalized across healthcare systems and improved further with local fine-tuning. Integrated explainability traced each risk update to patient-specific clinical factors, revealing distinct temporal persistence of prognostic information, with relevance half-lives ranging from weeks for therapies to nearly one year for baseline characteristics.

These findings demonstrate that learning from hospital-wide patient trajectories enables interpretable and continuously updated predictions, providing a scalable framework to support individualized treatment decisions.

## Introduction

Advances in cancer diagnosis and treatment have substantially improved patient outcomes at the population level. Yet, for individual patients, treatment outcomes remain highly variable and difficult to predict^1^. Each patient follows a unique clinical trajectory, shaped by individual tumor biology, host factors, comorbidities, treatment interventions, and pathophysiological responses that need to be assessed continuously to guide personalized care. However, despite the wealth of longitudinal data generated throughout a patient journey, clinical decision-making relies predominantly on population-level evidence and physician experience.

Prognostic models have long sought to address this gap. Traditional scoring systems and, more recently, artificial intelligence (AI) models have demonstrated promising predictive performance across a range of outcomes ^2–8^. In our own prior work, we have shown how large-scale real-world data can be leveraged by explainable AI to inform pan-cancer treatment guidance^3,7^. Building on this foundation, a remaining key challenge for the widespread application of such models is that they typically represent cross-sectional data, disregarding both the full longitudinal trajectory of patients and the heterogeneous nature of real-world data ^3,8–11^. Clinical translation instead requires interpretable models that take into account individual disease trajectories across patient populations and adapt flexibly to complete electronic health records (EHR), handling temporality and irregular or missing data points while remaining customizable to predict actionable clinical outcomes. Transformer models, originally developed for natural language processing, are well suited to this setting, with their capacity to model complex sequential data. Initial applications to longitudinal EHR are emerging, but existing approaches capture only a narrow subset of clinical information, provide insufficient transparency, or have uncertain generalizability^4,12,13^. This leaves open whether the dense information stream of routine oncological care can be modeled to provide continuously updated, individualized risk profiles that are clinically actionable and generalizable across hospitals.

Here, we present Chronicle, a transformer-based architecture that models entire cancer patient trajectories for explainable, multi-endpoint outcome prediction. Chronicle was trained on complete longitudinal records from 51,711 patients across 67 cancer types, integrating 562 clinical variables, spanning demographics, staging, laboratory values, vital signs, diagnoses, and therapies. Chronicle processes these sequences in their native form and generates dynamic, patient-level risk predictions that update continuously as new clinical information becomes available. It is customizable to diverse prediction tasks, including overall survival, adverse events, and transfusion needs, while the architecture extends to arbitrary additional endpoints. To facilitate clinical application, integrated layer-wise relevance propagation (LRP)^14–17^, adapted to transformer architectures, provides native explainability that traces how individual clinical events contribute to each predicted outcome over time. We validated Chronicle in a multicenter setting, spanning three external cohorts across three healthcare systems, examining both the prediction performance and the explained decision-making.

This work demonstrates the potential of explainable transformers trained on hospital-wide cancer records and establishes Chronicle as a scalable backbone for AI-informed precision oncology.

## Results

### Cohort description

We retrospectively assembled four independent pan-cancer cohorts with solid tumors from university hospitals in Germany, Switzerland, and the United States. The training cohort comprised 53.7 million data points from 51,711 patients across 67 cancer types treated at University Hospital Essen between January 2000 and November 2025 (**Fig. 1A**). Eligible patients were adults with a solid malignancy and an observation period containing a minimum number of recorded data points (see **Suppl. Fig. 1** for the patient inclusion process). The data encompassed 562 clinical variables, including demographics, tumor characteristics (including histology and TNM stage), laboratory measurements, systemic treatments, and clinical procedures. The data density across feature groups is visualized in **Supplementary Figure 2**. During follow-up, 16,859 patients (32.6%) died, and between 7.5% and 22% experienced at least one severe adverse event or required blood transfusion (**Table 1**).

**Figure 1:**
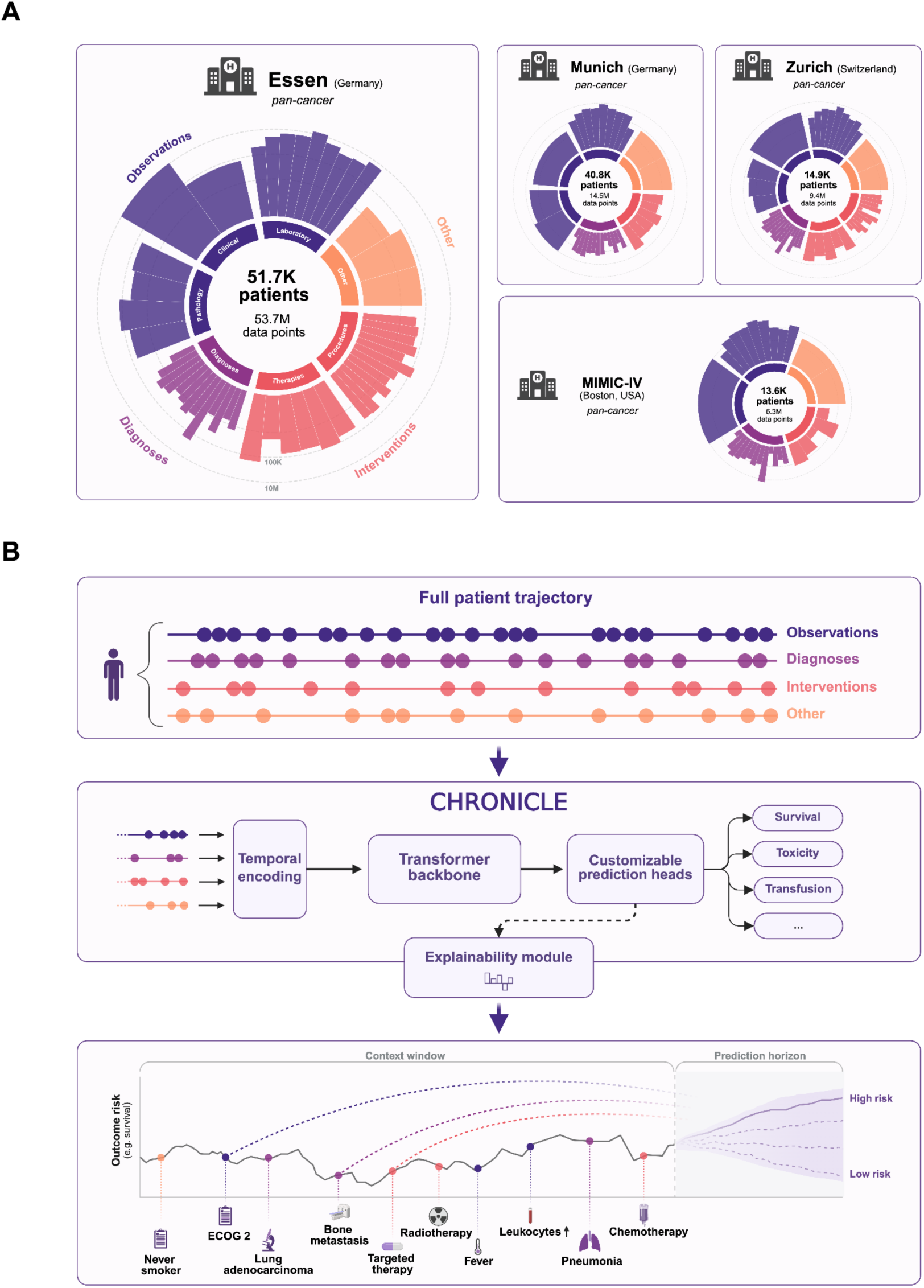
Patient cohorts and study design A: Data composition of pan-cancer patient cohorts from four university medical centers across three countries: Essen (containing the primary training cohort) and Munich (Germany), Zurich (Switzerland), and MIMIC-IV (Boston, USA). Inner-ring segments group variables into top-level categories (Observations, Diagnoses, Interventions, Other). Outer-ring bars show subgroups within each category, with height encoding the number of data points (log₁₀ scale). **B:** Chronicle prediction workflow. Full patient trajectories are temporally encoded and processed by a transformer backbone. Customizable prediction heads generate predictions for clinical endpoints of interest, with an integrated explainability module identifying the influence of all patient information in the model’s context window on individual predictions.

**Table 1:** Cohort description.

|  | **Essen**<br>(Germany) | **Munich**<br>(Germany) | **MIMIC-IV**<br>(Boston, USA) | **Zurich**<br>(Switzerland) |
| --- | --- | --- | --- | --- |
| **Patients, *n*** | 51,711 | 40,841 | 13,632 | 14,868 |
| **Data points, *n*** | 53,724,193 | 14,523,496 | 6,297,448 | 9,352,085 |
| **Variables, *n*** | 562 | 193 | 231 | 485 |
| **Sex, %** |  |  |  |  |
| Female | 45.0 | 43.9 | 46.1 | 39.5 |
| Male | 55.0 | 56.1 | 53.9 | 60.5 |
| **Age, years** |  |  |  |  |
| Median | 63.7 | 64.0 | 68.0 | 64.5 |
| Range | 18.0 - 106.4 | 18.0 - 99.0 | 18.0 - 99.0 | 18.1 - 99.1 |
| **Record span (days), median<sup>a</sup>** | 416<br>(407.0-425.0) | 226<br>(219.0-233.0) | 92<br>(83.5-101.0) | 595<br>(575.0-614.0) |
| **Deceased, %** | 32.6 | 21.5 | 37.8 | 41.8 |
| **Adverse events, *n* (%)** |  |  |  |  |
| Anemia<sup>b</sup> | 11,384 (22.0%) | 6,703 (16.4%) | 3,710 (27.2%) | 3,509 (23.6%) |
| Neutropenia<sup>b</sup> | 5,568 (10.8%) | 1,017 (2.5%) | 634 (4.7%) | 2,011 (13.5%) |
| Thrombocytopenia<sup>b</sup> | 3,879 (7.5%) | 1,482 (3.6%) | 1,010 (7.4%) | 1,077 (7.2%) |
| Elevated ALT<sup>c</sup> | 7,889 (15.3%) | 4,359 (10.7%) | 1,884 (13.8%) | 2,352 (15.8%) |
| Elevated AST<sup>c</sup> | 8,627 (16.7%) | 1,970 (4.8%) | 2,101 (15.4%) | 2,115 (14.2%) |
| Elevated creatinine<sup>c</sup> | 6,139 (11.9%) | 3,850 (9.4%) | 2,455 (18.0%) | 3,325 (22.4%) |
| **Received blood transfusion, *n* (%)** | 9,459 (18.3%) | Not available | 1,428 (10.5%) | Not available |

<sup>a</sup> Defined as the time between the first and last available record per patient; values in parentheses denote the 95% confidence interval of the median, estimated via bootstrap resampling (2,000 resamples).
<sup>b</sup> CTCAE grade $\geq 3$ .
<sup>c</sup> Elevated above upper limit of normal (ULN): $>3\times$ ULN for ALT (alanine aminotransferase) and AST (aspartate aminotransferase), $>1.5\times$ ULN for creatinine, based on CTCAE grade $\geq 2$ thresholds.

To assess generalizability across hospital settings, we evaluated the model on three external cohorts from the University Hospital Zurich (USZ), LMU University Hospital Munich, and Beth Israel Deaconess Medical Center (MIMIC-IV)^18,19^, encompassing 69,341 patients and 30.2 million data points in total. Detailed cohort characteristics are provided in **Table 1**. The distribution of primary tumor sites for all cohorts is shown in **Supplementary Figure 3**.

### Chronicle learns entire treatment trajectories for multiple outcome prediction

We developed Chronicle, a transformer-based framework that encodes longitudinal patient trajectories as sequences of clinical events and continuously updates its internal representation over time. This design reflects the reality of clinical practice, where data accumulates irregularly and decisions must be made with whatever information is available. Unlike conventional models that rely on data collected at fixed timepoints (cross-sectional data), Chronicle natively handles irregular and missing data without imputation, operating directly on timestamped clinical records from routine care.

Trained on the Essen cohort using a multitask objective, Chronicle can predict a wide range of clinical outcomes. In this study, we chose eight clinically relevant endpoints: overall survival, nadir levels of hemoglobin, neutrophils, and platelets within a 60-day window as indicators of hematological toxicity, liver injury through elevated levels of alanine aminotransferase (ALT), aspartate aminotransferase (AST), and kidney function through creatinine within a 60-day window, defined relative to sex-specific upper limits of normal, and administration of blood transfusions occurrence within a seven-day horizon.

A schematic overview of the model architecture and training procedure is provided in **Fig. 1B**.

### Accurate prediction of overall survival and adverse events along patient trajectories

Accurate prediction of survival and adverse events is central to personalized cancer management, as it enables timely intervention and informed treatment decisions. Chronicle achieved a concordance index (C-index) of 0.84 for overall survival prediction on a held-out test set from the internal Essen cohort, outperforming Cox proportional hazards models (C-index: 0.76, P<0.001), neural networks (0.77, P<0.001), and random survival forests (0.79, P<0.001) **(Fig. 2A)**. Notably, whereas the cross-sectional baseline models required imputation and additional preprocessing to handle missing and irregularly sampled values, Chronicle operated directly on the raw clinical records, preserving the underlying data structure.

**Figure 2:**
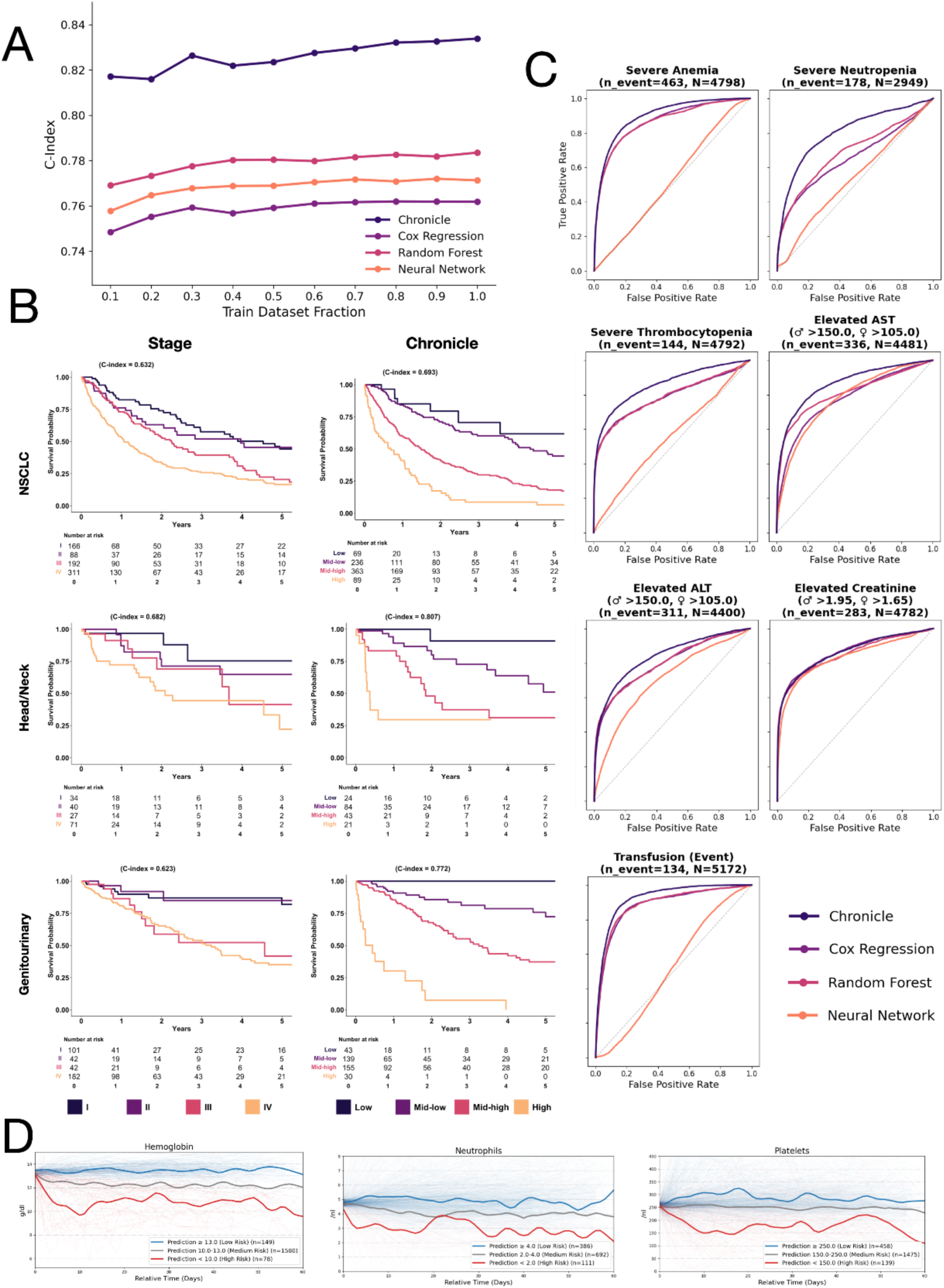
Predictive performance of Chronicle across survival and adverse event tasks. **A**: Scaling study assessing model performance for the prediction of overall survival with respect to training dataset size. Models were trained on increasing fractions of the dataset (10% to 100%). **B**: Kaplan–Meier overall survival across three cancer cohorts (NSCLC, head and neck, genitourinary), stratified by Chronicle risk group (Low-High) and clinical TNM stage (I-IV), evaluated on the subset of test patients with available stage information over five years of follow-up, with patients at risk tabulated below each panel. Per-panel C-indices quantify discrimination of the respective stratifier, with higher risk scores and stages corresponding to shorter survival. **C**: Receiver operating characteristic (ROC) curves for the prediction of clinically relevant adverse events: severe anemia, neutropenia, thrombocytopenia (within 60 days), as well as elevated AST, ALT, creatinine (within 60 days), and blood transfusion (within 7 days). **D**: Lab value trajectories following a normal-range baseline. Patients were included if their real lab value fell within a predefined normal range at a given timepoint (baseline, t=0) and were stratified into risk groups based on the model’s prediction at that point. Individual patient trajectories over the following 60 days are shown as thin lines, with LOWESS-smoothed group medians overlaid (blue = low risk, grey = medium risk, red = high risk). Analysis was restricted to labels for which all risk groups contained more than 50 patients (hemoglobin, neutrophils, platelets).

Performance gains increased with dataset size, with Chronicle showing substantially greater improvement than single time-point models at larger cohort sizes (**Fig. 2A**), indicating that the longitudinal architecture benefits more strongly from larger training sets. Conducting an ablation study on the other patient outcomes showed a similar improvement in prediction performance with higher sample size for neutropenia and kidney function, while increased sample size showed less benefit for the remaining labels (**Suppl. Fig. 4**).

When stratified by tumor entity, Chronicle outperformed all baselines in 14 of 15 cancer types (except Skin NOS) with more than 100 patients in the test set (**Suppl. Fig. 5**), demonstrating consistent performance across heterogeneous patient populations. Chronicle also substantially outperformed the modified Glasgow Prognostic Score (mGPS; C-index 0.81 vs 0.58), derived from albumin and C-reactive protein (CRP) in patients with available data (**Suppl. Fig. 6**).

Chronicle further stratified patients with available staging information into distinct risk groups across three cohorts (NSCLC, head/neck, and genitourinary). Stratification by Chronicle risk group outperformed clinical TNM stage in discriminating overall survival in all three cohorts (C-index: NSCLC 0.693 vs. 0.632; head/neck 0.807 vs. 0.682; genitourinary 0.772 vs. 0.623; **Fig. 2B**), indicating that Chronicle captured prognostic variation not fully explained by stage alone.

The corresponding patient-level redistribution between stage and risk group is shown in Supplementary Figure 7.

Beyond the prediction of overall survival, Chronicle outperformed all baseline models across six of seven clinical event prediction tasks and performed comparably to the best baseline for elevated creatinine (**Fig. 2C).** Chronicle achieved AUROCs of 0.92 for blood transfusion, 0.90 for anemia, 0.86 for thrombocytopenia, 0.80 for neutropenia, 0.86 for elevated AST, 0.84 for elevated ALT, 0.86 for elevated creatinine, with improvements of up to 0.11 over the best baseline model (**Fig. 2C**).

To illustrate the potential clinical utility of predictions at the individual patient level, we examined longitudinal laboratory trajectories among patients with comparable baseline values, assessing whether Chronicle could anticipate divergent future trajectories despite similar starting points. Grouping these patients by their predicted hemoglobin, neutrophil, and platelet ranges, those in the lowest predicted range showed marked declines in the corresponding laboratory values over the subsequent 60 days, whereas those in the highest predicted range remained largely stable (**Fig. 2D**).

### Chronicle generalizes across independent healthcare systems

A major barrier to clinical deployment of AI models is their limited ability to generalize beyond the institution where they were developed.^20^ Differences in patient populations, clinical focus areas, diagnostic protocols, treatment practices, and documentation procedures between hospitals can substantially degrade model performance and limit real-world applicability.

To assess cross-institutional robustness across different countries, we evaluated the model on three independent external cohorts. Chronicle, trained on the Essen cohort, achieved a C-index of 0.84 on internal evaluation. When applied zero-shot, i.e. without fine-tuning, to three independent external cohorts, the model maintained strong performance for overall survival prediction with C-indices of 0.84, 0.72, and 0.80 for the Munich, Boston, and Zurich cohorts, respectively (**Fig. 3A**). This lower baseline performance in the MIMIC-IV cohort likely stems from the absence of TNM staging information in the dataset, presenting a more challenging prediction task. The overall results demonstrated robust transferability across institutions and countries. Generalization was strong for the prediction of severe anemia (0.87-0.89), thrombocytopenia (0.86-0.90), and neutropenia (0.79-0.84) across all three external cohorts. Prediction of further adverse events was similarly robust, with AUROCs of 0.83-0.85 for elevated AST, 0.83-0.84 for elevated ALT, and 0.87-0.90 for elevated creatinine across all three external cohorts. For the prediction of blood transfusion, performance remained robust at the MIMIC-IV cohort (AUROC 0.82), the only external cohort with available transfusion labels.

**Figure 3:**
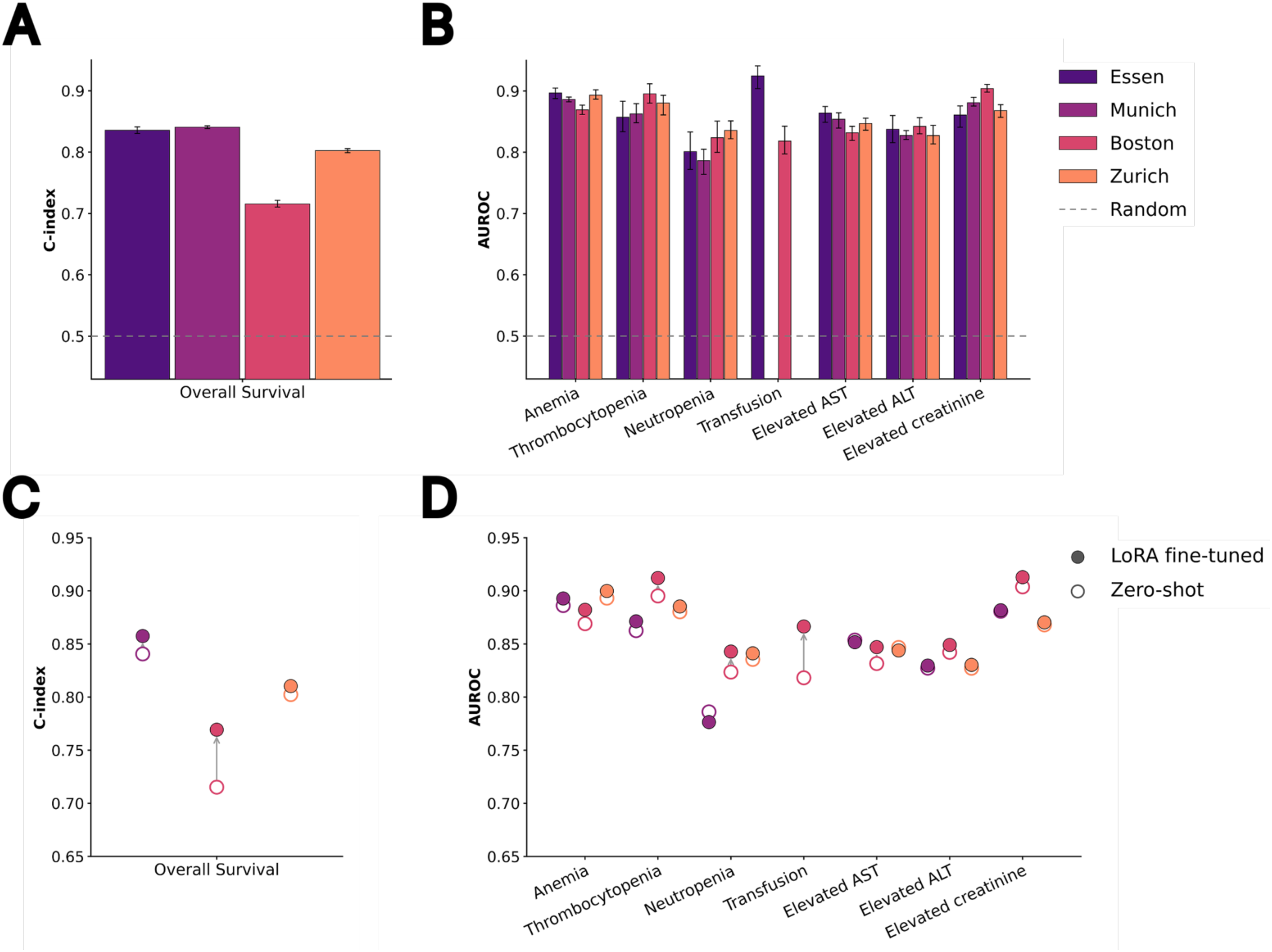
Chronicle discrimination performance and the effect of site-specific fine-tuning on external cohorts. A: C-index for overall survival prediction on the test set of the internal cohort (Essen) and three external validation cohorts (Munich, Boston, Zurich) under zero-shot transfer with frozen Essen-trained weights. **B:** AUROC for seven clinical endpoints. CTCAE grade ≥3 endpoints (anemia, thrombocytopenia, neutropenia) within 60 days, elevated AST and ALT (>3× ULN, analog to CTCAE v6) and creatinine (>1.5× ULN, analog to CTCAE v6) within 60 days and for transfusion within 7 days; transfusion data were unavailable for Munich and Zurich. **C, D:** Effect of site-specific LoRA fine-tuning of Chronicle on each external cohort (Munich, Boston, Zurich), compared against zero-shot transfer, for overall-survival C-index (**C**) and the cytopenia and transfusion endpoints (**D**). Fine-tuning was performed on a held-out training set for each respective site. Hollow circles indicate zero-shot performance, filled circles indicate LoRA fine-tuned performance, and arrows show the direction of change. In **A** and **B**, bars represent the mean, and error bars indicate the 2.5th–97.5th percentiles across 50 repeated evaluations, each based on one randomly selected prediction time point per patient. Essen results were computed with the same procedure on its held-out internal test set. The dashed line marks chance performance (0.5). In **C** and **D**, points are means over the same sampled points.

These results demonstrate robust cross-institutional generalization of Chronicle, supporting its use for clinical risk stratification across heterogeneous healthcare settings.

### Site-specific fine-tuning further improves performance across institutions

While Chronicle demonstrated already strong zero-shot generalization, its application in different clinical settings may further benefit from targeted adaptation to local data characteristics. This is particularly relevant for smaller institutions where training a model from scratch is not feasible.

To address this, we integrated an efficient adaptation of the low-rank adaptation (LoRA) into the Chronicle framework, allowing efficient fine-tuning on local data^21^. LoRA fine-tuning was applied across all three external cohorts on a held-out fine-tuning dataset (15% of data), mimicking the setting of a small institute’s limited data resources. Performance on overall survival prediction improved across all three cohorts, with the largest gain on MIMIC-IV (0.72 to 0.77), followed by Munich (0.84 to 0.86) and Zurich (0.80 to 0.81). Fine-tuning largely maintained or improved AUROCs for adverse events across all sites, with the largest and most consistent gains at Boston, spanning blood counts (neutropenia 0.82 to 0.84), blood transfusion (0.82 to 0.87), and liver and kidney lab test abnormalities (elevated creatinine 0.90 to 0.91, with elevated AST improving from 0.83 to 0.85 and ALT from 0.84 to 0.85). Improvements at Zurich were modest, with minor variation across endpoints (e.g., elevated AST 0.85 to 0.84), while performance at Munich was already high under zero-shot transfer, with AUROC endpoints remaining essentially unchanged despite a modest gain in overall C-index (0.84 to 0.86) (Fig. 3D). These results demonstrate that Chronicle can serve as a pre-trained backbone that can be efficiently adapted to new clinical environments with limited local data, facilitating deployment across diverse healthcare settings.

### Patient-level explainability enables continuous clinical interpretation

For clinical deployment, it is essential that model predictions can be interpreted in the context of individual patient trajectories. We therefore integrated LRP^11,16,22^, adapted for transformer architectures, in the Chronicle framework to quantify the contribution of each clinical event to the prediction over time. Because Chronicle recalculates its predictions each time new clinical data is recorded, both the risk and the events driving it are updated at every step. In routine use, a clinician would therefore not see a static score, but a continuously updated risk alongside the specific signals behind each change, mirroring how information becomes available over the course of care.

We illustrate this approach in a representative 60-year-old male patient with NSCLC adenocarcinoma (initial stage III) who underwent multimodal therapy, including radiotherapy and multiple lines of chemotherapy, immune checkpoint inhibitors, and antiangiogenic therapy. Over the disease course, the patient developed metastatic progression and multiple complications, including pulmonary embolism and pleural effusion, as well as hematologic toxicity requiring repeated blood transfusions and filgrastim support, before ultimately dying approximately four years after diagnosis **(Fig. 4A)**. Chronicle tracked the patient trajectory and continuously provided predictions for prognosis and adverse events. Predicted mortality risk remained low during the initial treatment phase before increasing progressively during later disease progression **(Fig. 4B)**. At day 535, the first marked increase in predicted risk was primarily attributed to the emergence of additional metastatic sites, together with reduced hemoglobin levels.

**Figure 4:**
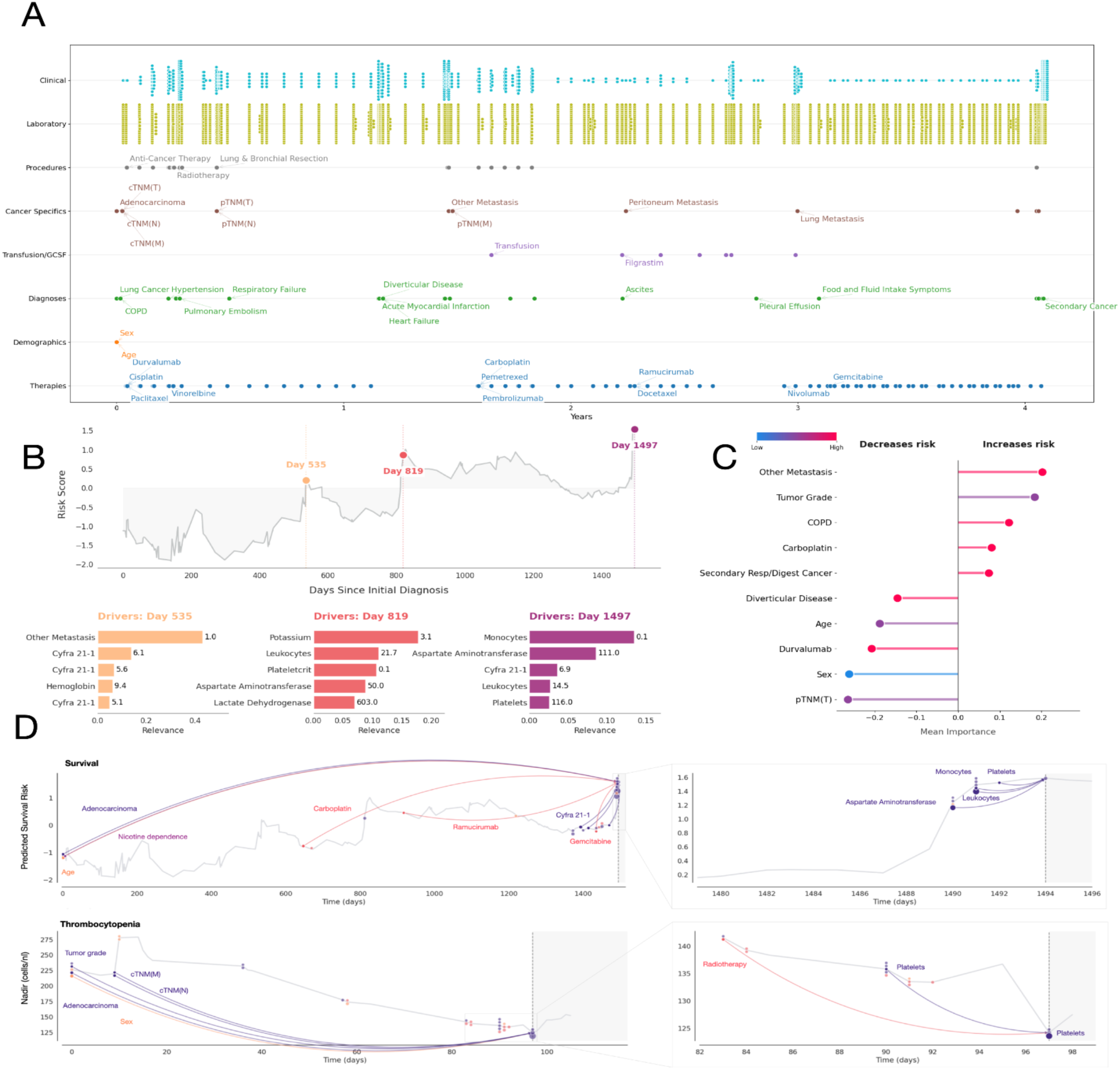
Comprehensive longitudinal analysis of a single patient with model predictions and LRP-based explanations. **A**: Timeline depicting the complete clinical trajectory from initial lung cancer diagnosis to death, structured by feature entities. The visualization captures disease evolution, therapeutic interventions, metastatic progression, and the occurrence of secondary malignancies, of which selected examples are shown. **B:** The line plot illustrates the predicted risk score (y-axis) over time since initial diagnosis (x-axis) for a representative patient. Three time points (Day 535, 819, and 1497) are highlighted. Below, bar charts for each selected time point display the five variables with the highest aggregated LRP scores, where the x-axis represents relevance magnitude and the y-axis lists the corresponding variables. **C:** A dot plot depicting mean LRP importance (x-axis) of the strongest positive and negative contributions to mortality risk prediction for the selected patient, color-coded by variable value (blue: low, red: high). Variables are ordered by their contribution to decreasing (left) or increasing (right) predicted risk. **D:** Longitudinal LRP-based explanations for two clinical outcomes (survival and 60-day nadir of platelets). For both outcomes, the left panel shows the full prediction trajectory until the explained timepoint (dashed vertical line) (x-axis: time in days; y-axis: predicted value or risk score), with the solid line representing the time of model prediction. Colored dots represent the top 30 patient characteristics that contribute most to the prediction. The right panel shows a zoomed view of the region immediately preceding the explained timepoint, highlighting the short-term temporal relevance pattern. Color coding distinguishes variables into four different groups (Observations, Diagnoses, Interventions, Other). While recent patient information has a dominant effect on the prediction, the model also relies on long-distance dependencies for patient characteristics such as age and TNM staging.

Using the built-in explainability, Chronicle identified Cyfra 21-1 as a relevant contributor across multiple consecutive measurements. The subsequent risk increase at day 819 was attributed to low potassium as well as high leukocyte levels, accompanied by alterations in plateletcrit, AST, and lactate dehydrogenase, indicating a broader inflammatory and metabolic disturbance alongside possible hepatic involvement. At the latest timepoint, day 1497, monocyte counts and

AST emerged as the strongest contributors to predicted mortality risk, together with Cyfra 21-1, leukocytes, and platelets.

Taken together, these findings show that Chronicle’s predicted risk integrates a broad and clinically diverse evidence base, capturing hematologic abnormalities, tumor-associated markers such as Cyfra 21-1, and additional organ-specific pathologies such as hepatic dysfunction, rather than relying on isolated acute events or a single category of clinical signal **(Fig. 4B)**. Aggregated patient-level feature attribution further demonstrated that metastatic disease characteristics, including metastases, high tumor grade, and COPD, were among the strongest contributors to increased predicted mortality risk in this patient **(Fig. 4C)**. In contrast, younger age at diagnosis and lower TNM(T) stage contributed favorably to the prognosis. Results for the remaining labels are shown in **Supplementary Figure 8**.

Chronicle provides separate explanations for each prediction task, allowing the temporal structure of the attributions to be compared across endpoints. We observed that Chronicle draws on distinct subsets of variables for each task, incorporating both short- and long-term dependencies (**Fig. 4D + Suppl. Fig. 9)**. For survival prediction, disease-related variables such as histological subtype (adenocarcinoma), nicotine dependence, and age contributed to the prediction across the entire context window, reflecting stable background risk, while treatment exposures (carboplatin, ramucirumab, gemcitabine) and proximal laboratory markers such as Cyfra 21-1, AST, monocyte, and leukocyte counts became dominant only immediately preceding the explained timepoint. Predictions of the minimum platelet nadir showed a comparable structure, drawing on temporally distant disease characteristics, including tumor grade, TNM stage, histological subtype, and sex, alongside proximal signals such as recent radiotherapy and preceding platelet trends shortly before the prediction horizon. Similar patterns with varying proportions of short- and long-term dependencies were observed for the remaining prediction tasks (**Suppl. Fig. 9)**. The consistent co-occurrence of distant and recent relevant variables across endpoints indicates that Chronicle learned to integrate long-range disease history with immediate clinical dynamics, assigning temporal relevance in a manner that is consistent with clinical oncologic reasoning. Together, this illustrates how Chronicle can track patient trajectories through the disease course, providing a continuously updated risk assessment across multiple outcome dimensions, with granular explanations for each endpoint.

### Cohort-level explainability identifies time-dependent effects of systematic prognostic drivers

To characterize which variables contributed most to Chronicle’s predictions overall, we calculated mean absolute LRP relevance scores across all patients in the Essen cohort. Among variables recorded in at least 50% of patients, neutrophils received the highest relevance for overall survival, followed by AST, hemoglobin, age, and ALT (**Fig. 5A**). These findings are consistent with the established central role of systemic inflammation, hematological status, and hepatic function in patients with cancer^23–26^. Further highly ranked variables were creatinine, metastatic status, and albumin, indicating that Chronicle relies on a multi-system characterization of the patient, including organ function, disease burden, and nutritional status, rather than on a single domain.

**Figure 5:**
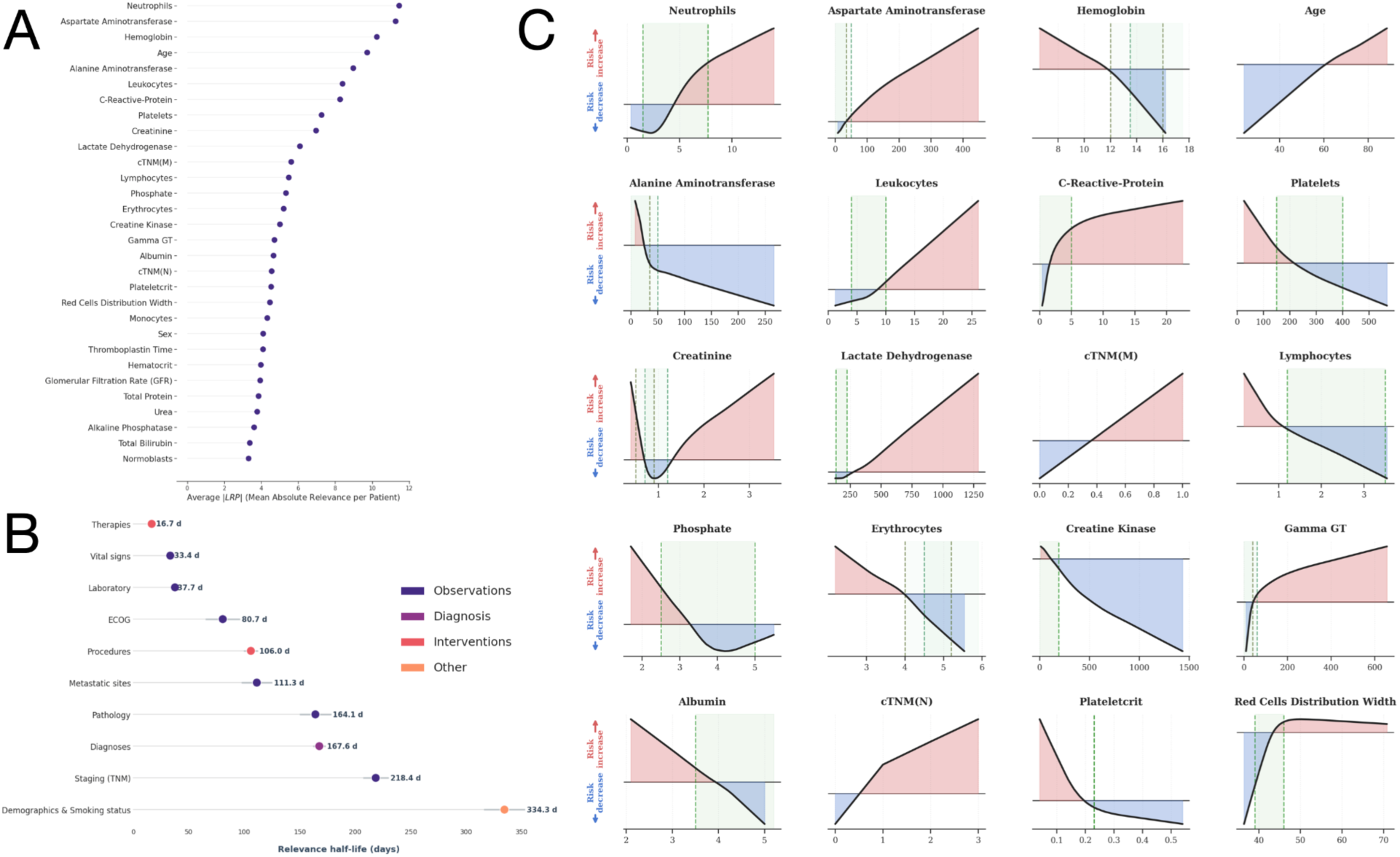
Analysis of variable-specific risk contributions, global importance, and relevance decay over time. **A**: A dot plot representing the mean absolute variable importance across the entire dataset of variables occurring in at least 50% of the patients. Variables are ranked by their average absolute LRP relevance. **B**: Analysis of variable relevance decay, showing the temporal stability of information across different feature groups. The x-axis denotes the number of days until the relevance of a measurement decreases by 50%. The plot displays the mean (dot) and the 95% confidence interval (error bars). **C**: Visualization of the relationship between individual variable values (x-axis) and their corresponding mean LRP-based risk contributions (y-axis). Positive values (red) indicate a higher than average predicted risk, while negative values (blue) indicate a lower than average predicted risk. The physiological reference ranges for each laboratory parameter are highlighted by green dashed vertical lines and shaded areas; when sex-specific differences exist, the reference ranges are shown separately for female (light green) and male (dark green) patients.

As Chronicle leverages the complete patient history rather than only the most recent clinical information, LRP enabled us to quantify how the relevance assigned to records evolved over time^27^. As expected, Chronicle placed overall greater emphasis on recent measurements than on those further in the past. This temporal decay, however, varied strongly between variable categories (**Fig. 5B**). Therapies showed the shortest relevance half-life (16.7 days), followed by vital signs (33.4 days) and laboratory values (37.7 days), consistent with the rapid dynamics of acute clinical changes. ECOG performance status remained relevant over a longer period (80.7 days), whereas procedures, pathology, diagnoses, and TNM stage decayed more slowly (106.0-218.4 days). Demographics (sex and age) and smoking status remained informative for the longest duration (mean 334.3 days), consistent with their role as static prognostic factors. These decay profiles provide a data-driven perspective on how long different clinical information retain predictive relevance and may inform the prioritization of monitoring strategies.

For the prediction of adverse treatment events, patient characteristics showed different patterns of relevance decline over time. (**Suppl. Fig. 10**). Overall, features stayed relevant longer for survival than for hematological outcomes (half-life 182.9 vs 134.1 days). Laboratory values showed a relevance half-life between 25.6 and 31.5 days for hematological adverse event prediction. The relevance of therapies had a half-life of 20.4 days for prediction of anemia but only 9.8 days for the prediction of thrombocytopenia, in line with the fast regeneration of platelets, while vital signs had a similar relevance half-life for all adverse events (26.3-32.5 days).

Next, we examined how Chronicle assigned risk across the value range of individual variables. Variable-level attribution profiles were broadly consistent with established clinical knowledge, with the direction of risk assignment matching clinical expectations across all predicted outcomes (**Fig. 5C**). For example, higher inflammatory markers, such as leukocyte counts and C-reactive protein, were associated with increased contribution to mortality risk. However, the attribution profiles differed between markers: whereas leukocyte counts showed a marked increase in risk contribution only above the normal range, even modest increases in CRP contributed to higher predicted mortality risk within the clinically normal range.

In contrast, for other variables, including platelets, erythrocytes, and hemoglobin, risk-increasing contributions emerged below their respective reference ranges. Several variables, including neutrophil count, creatinine, and phosphate, exhibited non-monotonic attribution profiles, with increased risk contributions at both low and high values and a minimum within the reference range. Such patterns cannot be represented by models that assume linear covariate effects, illustrating the benefit of flexible modeling approaches for identifying complex risk patterns.

For some variables, including hemoglobin and creatinine, the transition from risk-decreasing to risk-increasing contributions closely coincided with established clinical thresholds. For others, including CRP, neutrophil counts, and phosphate, these transition points deviated from conventional reference limits, suggesting that the value ranges Chronicle associated with elevated risk are not uniformly captured by standard clinical dichotomizations. Notably, both low and high creatinine values were assigned risk-increasing contributions. Low creatinine may reflect reduced muscle mass in the context of cachexia and frailty in patients with cancer, whereas elevated creatinine is consistent with impaired renal function^28,29^. Conversely, high creatine kinase (CK) values contributed to lower risk. Given the high prevalence of sarcopenia in patients with cancer, elevated CK may serve as a surrogate marker of preserved muscle mass, potentially explaining its association with lower predicted risk^30^. The trained model showed highly consistent attribution profiles when applied to a random subset of the external Munich cohort (**Suppl. Fig. 11**).

## Discussion

Cancer treatment consists of sequential interventions whose effects accumulate over time, yet most models developed to support oncological decisions represent patients at a single timepoint. Here we presented Chronicle, a transformer-based model trained on longitudinal clinical records from 51,711 patients across 67 cancer types that simultaneously predicts survival and seven treatment-associated adverse events using routinely collected clinical data. Chronicle achieved strong prediction performance across tasks, generalized to independent external cohorts, and provided patient-level interpretability, addressing several barriers that have limited the clinical translation of AI in oncology.

Existing prognostic models are typically developed for specific cancer types and rely on data collected at fixed timepoints, limiting their applicability to the heterogeneous and incomplete records encountered in routine care^10,31^. Chronicle overcomes these constraints by directly modeling irregular longitudinal patient trajectories without requiring explicit handling of missing values through imputation, thereby preserving the original temporal structure of clinical data while avoiding the introduction of systematic biases. The consistent improvements over single-time- point models indicate that longitudinal modeling captures evolving disease trajectories and early clinical signals that remain inaccessible to static approaches. For overall survival prediction, Chronicle showed continued performance gains as training cohort size increased, consistent with scaling behavior observed in large language and vision models^32,33^. These improvements exceeded those of conventional machine learning models across increasing cohort sizes. ^4,32,34^. Moreover, Chronicle maintained robust performance even on datasets containing only subsets of variables, consistent with the extensive data augmentation during training and suggesting that pretrained longitudinal models may retain utility even in data-constrained settings.

Beyond predictive performance, Chronicle provides temporally resolved explanations of individual patient trajectories that can be calculated in real time. Both patient- and cohort-level analyses identified established prognostic factors, including age, C-reactive protein, and TNM stage, supporting the clinical plausibility of Chronicle’s decision-making^26^. Temporal analyses further revealed distinct half-lives of predictive relevance across variable categories, with rapidly changing variables such as laboratory values and medications requiring frequent reassessment, whereas demographics and staging remained informative over longer periods. These temporal relevance patterns varied across prediction tasks, with hematological adverse events relying predominantly on more recent clinical information than overall survival, consistent with the faster dynamics of treatment-related toxicities compared with long-term prognosis. These findings provide a data-driven perspective on the temporal relevance of clinical information and may inform prioritization of monitoring strategies in routine care.

Chronicle generalized to cohorts from Germany, Switzerland, and the United States without retraining, demonstrating robustness to variation in healthcare systems, documentation practices, and coding standards. Given the substantial heterogeneity of real-world clinical data, this robustness suggests that large-scale longitudinal training may confer resilience to distributional shifts that often limit the deployment of task-specific models^35,36^. Together with efficient site-specific fine-tuning via LoRA, these findings support a practical deployment paradigm in which a shared pre-trained model is distributed across institutions and refined using limited local data.

Among the predicted outcomes, adverse events appear particularly well suited for early clinical implementation. Predicted adverse events can be met with supportive measures already established in routine oncological care, including intensified laboratory monitoring, growth-factor support, and transfusion planning. These interventions are widely available and generally carry limited risk, making adverse-event prediction an attractive first application for prospective evaluation. Individualized prognostic predictions hold considerable potential, but their interpretation and translation into clinical decisions pose additional challenges that warrant further investigation.

Several limitations should be considered. Chronicle was trained on observational data, in which treatment allocation was not randomized, precluding causal interpretation of variable–outcome relationships^37,38^. Correlations among clinical variables may enhance predictive performance but complicate attribution, as relevance can be distributed across collinear variables. Although Chronicle demonstrated strong external generalization, performance may vary in settings that diverge further from the training distribution. Finally, Chronicle currently operates exclusively on structured clinical data and does not yet incorporate imaging or text modalities, which may further enhance predictive performance and biological interpretability^8,39–41^.

Collectively, these findings show that transformer models trained on hospital-wide longitudinal cancer records can predict diverse clinical outcomes, generalize across healthcare systems, and provide interpretable, patient-level insights from routinely collected clinical data. More broadly, these results indicate that much of the information required to anticipate the clinical course is already contained in routinely generated healthcare data, distributed across time in a learnable structure. Chronicle could therefore complement recent large language model agents, which operate at the level of clinical reasoning rather than quantitative prediction, by supplying patient-specific risk profiles that these agents could act on within clinician-led workflows^42,43^. Extending Chronicle to additional data modalities and clinical applications may thus contribute to scalable frameworks for personalized decision support across diverse healthcare settings. Carefully designed prospective studies will be essential to determine appropriate use cases, establish decision thresholds, and evaluate whether such AI-supported approaches improve patient outcomes and quality of care.

## Methods

### Study design and data acquisition

We used the Smart Hospital Information Platform (SHIP) of University Hospital Essen to generate the internal dataset^44^. We identified all patients with solid cancers treated at the hospital between January 2000 and November 2025. Patients with hematologic malignancies were not considered. Patients were excluded if they were under 18 years of age, or if fewer than 50 data points were recorded across all variables. The final internal dataset comprised 51,711 patients. The detailed patient inclusion process is provided in **Suppl. Fig. 1**. Overall survival was defined as the time from the day of prediction to death from any cause. Dates of death were retrieved from medical records or, where unavailable, from the state cancer registry. Patients without a recorded date of death were right-censored on the date of their last clinical visit.

The study was approved by the Ethics Committee of the Medical Faculty of the University of Duisburg-Essen (22-10881-BO). The requirement for written informed consent was waived due to the study’s retrospective design and the deidentification of data. Data analysis in Zurich was approved by the Cantonal Ethics committee; BASEC-Nr: 2023-00234, “Comprehensive Cancer Center Zürich (CCCZ) - Data analytics”. The study was approved by the Ethics Committee at LMU Munich (23-0650).

### Data preprocessing

A two-step outlier removal approach was applied across numerical variables: In the first step, extreme outliers were manually curated. Subsequently, outlier values, defined as a deviation of more than three standard deviations from the mean, were removed. This method was specifically applied to laboratory values, clinical values, BMI, body surface area, and smoking pack-years.

Next, we performed variable-specific cleaning and standardization. Laboratory variables measured in fewer than 5,000 patients were excluded. Manual data cleaning was conducted using domain expertise to harmonize laboratory measurements. Clinically equivalent assays were merged into unified variables. For differential blood counts, only absolute cell counts were retained, whereas percentage-based values were excluded to reduce strong correlation between variables. This resulted in 110 unique laboratory variables.

Primary and secondary ICD-10 codes were truncated to the first three characters (e.g., C50.9 to C50). Primary ICD codes recorded in fewer than 10 patients were discarded. Secondary ICD codes were retained only if recorded in at least 500 patients. Each patient was assigned a primary ICD code, corresponding to a cancer diagnosis. The combination of primary and secondary ICD codes resulted in 277 unique codes.

Systemic cancer treatments were grouped into their common active ingredient. After removing duplicate patient-date records, we retained 35 unique substances and aggregated the 113 substances administered to fewer than 200 patients into a category “other”.

Prescription substances given to at least 200 patients were encoded as distinct variables, resulting in 14 distinct prescription variables. Substances given to fewer than 200 patients were grouped into “other_prescriptions” (61 prescriptions grouped).

German operation and procedure classification system (OPS) codes were truncated to the first three digits. Codes starting with ‘1’ or ‘3’ were excluded, and only codes occurring in at least 500 patients were retained. Duplicate entries for the same patient and date were excluded. After these filtering steps, 70 unique OPS codes remained.

TNM staging and ECOG Performance status were mapped to numerical scores. ICD and OPS codes and treatment information were encoded as a binary variable (0 = absent, 1 = present).

Metastasis codes were similarly encoded. The final dataset comprises 562 variables. To ensure consistency, all variable names were mapped to a standardized nomenclature using a global format file.

Lastly, outcome labels were generated. For laboratory-derived adverse event prediction, the nadir or peak of each laboratory variable within the following 60 days was used. For transfusion prediction, a binary indicator was defined as 1 if a transfusion occurred within the following 7 days and 0 otherwise. Laboratory-derived adverse events were informed by CTCAE v6.0. Severe anemia, neutropenia, and thrombocytopenia were defined according to grade ≥3 thresholds. Elevations in ALT, AST, and creatinine were based on grade ≥2 thresholds, defined relative to the fixed, sex-specific limits of normal rather than to individual baseline values for methodological consistency.

### External validation cohorts

External validation was performed on three independent cohorts: LMU University Hospital Munich (Germany), University Hospital Zurich (Switzerland), and the MIMIC-IV database (Beth Israel Deaconess Medical Center, Boston, USA). Patient selection followed the same criteria as in the internal Essen cohort, adapted to each site’s data capture period. All cohorts were preprocessed following the Essen pipeline described above, with site-specific adaptations to harmonize data structures, variable nomenclature, and units to the Essen-defined schema. After harmonization, 193 variables were retained for Munich, 485 for Zurich, and 231 for MIMIC-IV.

For Munich and Zurich, transfusion labels were unavailable. For MIMIC-IV, cancer cases were defined using ICD-9 and ICD-10 codes, with ICD-9 mapped to ICD-10 via the icdmappings library (https://github.com/snovaisg/ICD-Mappings).

### Model architecture

To capture the longitudinal nature of patient records, we implemented a longitudinal Transformer-based architecture using PyTorch^45^. The model processes complete patient trajectories, starting with foundational baseline information: for every patient, the primary tumor diagnosis, age, and sex are provided at Day 0 as the initial clinical context.

Before model training, each patient’s complete medical history was structured as a TensorDict^46^ (using keys variable, value, time, and all labels for each time point) capturing their entire multi-modal time-series trajectory. To account for different scales and distribution shapes, all continuous input variables as well as continuous prediction labels were normalized using the PowerTransformer (Yeo-Johnson transformation) from the *scikit-learn* library^47^. Test sets were normalized using the parameters fitted on the training set.

Each longitudinal measurement was represented as the sum of three component embeddings. These comprised a sinusoidal positional encoding of the time since initial diagnosis (days), a trainable embedding of the variable identity, and a linear projection of the measurement value. Additional [CLS] tokens were inserted for each day to provide a condensed daily summary of the patient’s state.

Sequences were processed by a Transformer encoder with an embedding dimension of 128, 8 attention heads, and 4 layers. To maintain computational efficiency, the maximum sequence length was capped at 4000 tokens during training. For patients exceeding this limit, variables were randomly subsampled while preserving baseline characteristics (primary tumor type, age, and sex) recorded at day 0.

A causal attention mask was used to enforce autoregressive attention. Outcome predictions were generated by multi-layer perceptron (MLP) heads with a hidden dimension of 256, mapping from [CLS] token representations. This modular design enables additional labels or clinical tasks to be incorporated by attaching further prediction heads. The resulting model comprised 2,345,862 trainable parameters.

We optimized overall survival prediction from right-censored labels using the negative Cox partial log-likelihood. Each patient contributes multiple CLS tokens, one per longitudinal time point. To avoid within-patient dependence between prediction timepoints in the loss, we randomly sampled a single risk score per patient at every training step and used it to compute the Cox likelihood loss. Validation followed the same per-step sampling scheme. To reduce variance from this stochastic selection, we repeated it ten times per validation epoch and reported the mean C-index. At test time, we drew 50 independent per-patient samples of time points, computed the C-index for each, and reported the mean as the final test performance.

Continuous labels were optimized using Mean Squared Error (MSE). Regression was used to predict hematological, renal, and hepatic adverse events^48^. A binary label for the endpoint transfusion requirement within 7 days was optimized using Binary Cross-Entropy (BCE) with Logits.

To optimize these tasks simultaneously, we employed Uncertainty Weighting^49^. To enhance the model’s ability to generalize, we implemented a dual-stage masking strategy:

Entire variable categories were masked with a probability of up to 20% (Feature-level Masking). Individual measurements were randomly masked with a probability of up to 20% (Data-point Masking).

We utilized a 90/10 split for training and testing. Additionally, 5% of the training dataset was held out as a validation dataset for the selection of hyperparameters, such as early stopping and learning rate scheduling. Models were trained using the Adam optimizer (learning rate: 10^-3^) with weight decay (10^-6^), gradient clipping, and a cosine annealing learning rate scheduler. Internal dropout was set to 0.1. Models were selected using early stopping based on the maximum C-index in the validation dataset with an early stopping patience of 20 epochs and a maximum number of epochs of 100. All experiments were conducted using the PyTorch framework on NVIDIA CUDA-enabled hardware.

### Benchmarking framework

To evaluate the predictive performance in a cross-sectional setting, we transformed the longitudinal dataset into a single cross-sectional representation. For each patient, one random time point was sampled from their clinical trajectory. At this sampled time point, the available clinical variables were extracted to form the input representation.

To mitigate the impact of missing data, a 7-day look-back window was applied. If a variable was not recorded at the selected time point, the most recent observation within the preceding seven days was carried forward. Remaining missing values were imputed using the *SimpleImputer* from scikit-learn^47^ with a median imputation strategy.

Variables were scaled using the PowerTransformer from scikit-learn, fitted exclusively on the training set. We implemented a multi-modal benchmarking suite across three label types: Survival (overall survival), which was evaluated using a Random Survival Forest, Cox Proportional Hazard model, and a Deep Neural Network with Cox Partial Likelihood loss; Binary Labels, which were evaluated using a Random Forest Classifier, Logistic Regression, and a Deep Neural Network (MLP) with Binary Cross-Entropy loss; and Continuous Labels, which were evaluated using a Random Forest Regressor, Elastic Net, and a Deep Neural Network with MSE loss.

To ensure a fair comparison, the longitudinal models were trained on the same patient cohorts as their cross-sectional counterparts. While cross-sectional models only observed the snapshot data available at the specific cutoff day (plus the 7-day look-back), the longitudinal models were granted access to the entire historical trajectory of the patient up until that point.

### Model explainability

We applied LRP to a transformer-based model for explanations of sequential predictions. LRP is an established method for explaining different model architectures ^16,17^, including transformer models ^50,51^.

For an input sequence *x*_1:*t*_ = (*x*_1_, …, *x_t_*), the model predicts the outcome sₜ at time t, i.e., *f*(*x*_1:*t*_) = *s_t_*. We aim to attribute the prediction *s_t_* to the input features in *x*_1:*t*_. Specifically, for each time step *t*, we compute relevance scores *R_t_*(*x_i_*) for all *i* = 1, …, *t*, where *R_t_*(*x_i_*) quantifies the contribution of input *x_i_* to *s_t_*ₜ

LRP uses layer-wise propagation rules that redistribute relevance from higher to lower layers. For each layer *l*, we specify a propagation rule 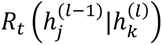 that assigns relevance from the k-th neuron in layer *l*, with activation 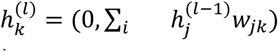, to the *j*-th neuron in layer (*l* − 1), with activation 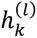. Our transformer model consists of feed-forward layers with ReLU activation, self-attention, and layer normalization. Specifying propagation rules for each component determines the relevance scores *R_t_*(*x_i_*).

For a feed-forward layer with ReLU activation, i.e., 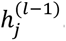, relevance is propagated recursively via the following rule (LRP-*γ* rule):

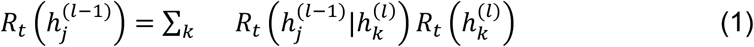

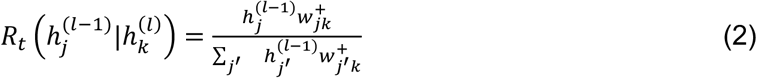

where *w*_j*k*_ are model parameters and *w*^+^ = *w*_j*k*_ + *γ*:0, *w*_j*k*_; with *γ* ≥ 0.

For self-attention and layer normalization, we use the LRP rules described in ^50^, which includes detaching the attention scores and the denominator of the layer normalization before applying LRP-*γ* rule. Cohort-level attribution profiles for the external Munich cohort were computed on a random subset of 10,000 patients drawn from the held-out Munich test set.

### Statistical analysis

All tests were two-sided and results were regarded as significant if *P* < 0.05. Kaplan-Meier plots were computed with the R package survival^52^. Predicted mortality risk was split into four groups by equidistant thresholds spanning the 2nd-98th percentile of the risk distribution within each tumor cohort with available staging information. C-Indices were calculated using lifelines^53^. AUC analyses were performed in scikit-learn47. Statistical tests between survival C-indices were calculated using the survcomp package54. Relevance half-life of a feature was defined as the shortest time between predictions for which the contribution of this feature decreased from peak relevance to ≤50% of its peak value.

### Visualizations

Figure 1 was created in BioRender: Kleesiek, J. (2026) https://BioRender.com/vp8xtar.

### Use of AI tools

The authors disclose the use of Claude (Anthropic) and ChatGPT (OpenAI) for language editing during the preparation of this article.

## Data Availability

Deidentified data are available upon reasonable request. Data cannot be shared with investigators outside the institution without consent. Data access is generally subject to applicable data protection regulations and data use and access regulation onsite. The MIMIC-IV database is available via PhysioNet to credentialed users (https://physionet.org/).

## Acknowledgments

The data from University Hospital Essen was provided by the Smart Hospital Information Platform (SHIP), managed by the Data Integration Center at the University Medicine Essen. SHIP serves as a comprehensive digital health platform for integrating data from all major clinical subsystems using a holistic FHIR-based approach. It enables the purification, analysis, distribution, and visualization of clinical data. Computations were carried out by KI Translation Essen (KITE). The data from University Hospital Munich was provided by Medical Data Integration Center at the LMU University Hospital (MeDIC^LMU^) via its Ecosystem for Digital Health Intelligence. It enables, empowers and orchestrates data-driven research and serves as a comprehensive platform for interoperable data integration, transformation, sharing as well as analysis and visualization in a trusted research environment. J.T.S. is supported by the German Cancer Consortium (DKTK) and the Deutsche Krebshilfe (DKH, German Cancer Aid) through 70117115 (ONCOverse).

## Contributions

Conceptualization: P.K and J.Keyl

Methodology: P.K., N.K., J.Bosserhoff, J.Keyl, T.L., T.S. Formal analysis: P.K., N.K., J.Bosserhoff, J.Keyl, T.N. Investigation: all authors.

Resources: B.F., S.S., S.Kuss, M.J.I., P.J., M.Kim, S.B., N.B., M.F., D.F.-S., M.Glas, V.G., B.H.,

K.H., S.Kasper, R.K., S.L., I.P., T.R., A.R., J.T.S., M.Guberina, U.S., M.Wichert, M.Ingrisch, K.U.,

J.Behr, D.T., C.G.S., J.M., N.H., A.Tufman, J.R., L.H.L., S.P., G.Höglinger, S.M., M.C., L.H., C.S.,

A.Tasdogan, M.T., A.W., M.Wiesweg, C.B.W., F.Albashiti, R.T., G.M., N.Mi., Z.B., M.v.B.-B., V.H.,

C.B., S.H., A.Wi., F.N., D.S., M.Krauthammer, K.-R.M., M.Schuler, F.K., J.Kleesiek Data acquisition: J.Bosserhoff, J.Keyl, P.K., N.K., T.L., T.N., F.Aubele Writing—original draft: P.K., N.K., J.Bosserhoff, J.Keyl, T.L.

Writing—review and editing: all authors. Supervision: J. Keyl, F.K., J.Kleesiek

