## Supplementary Material for "An explainable transformer model learning from entire treatment timelines for pan-cancer risk profiling across healthcare systems"

### Supplementary Information

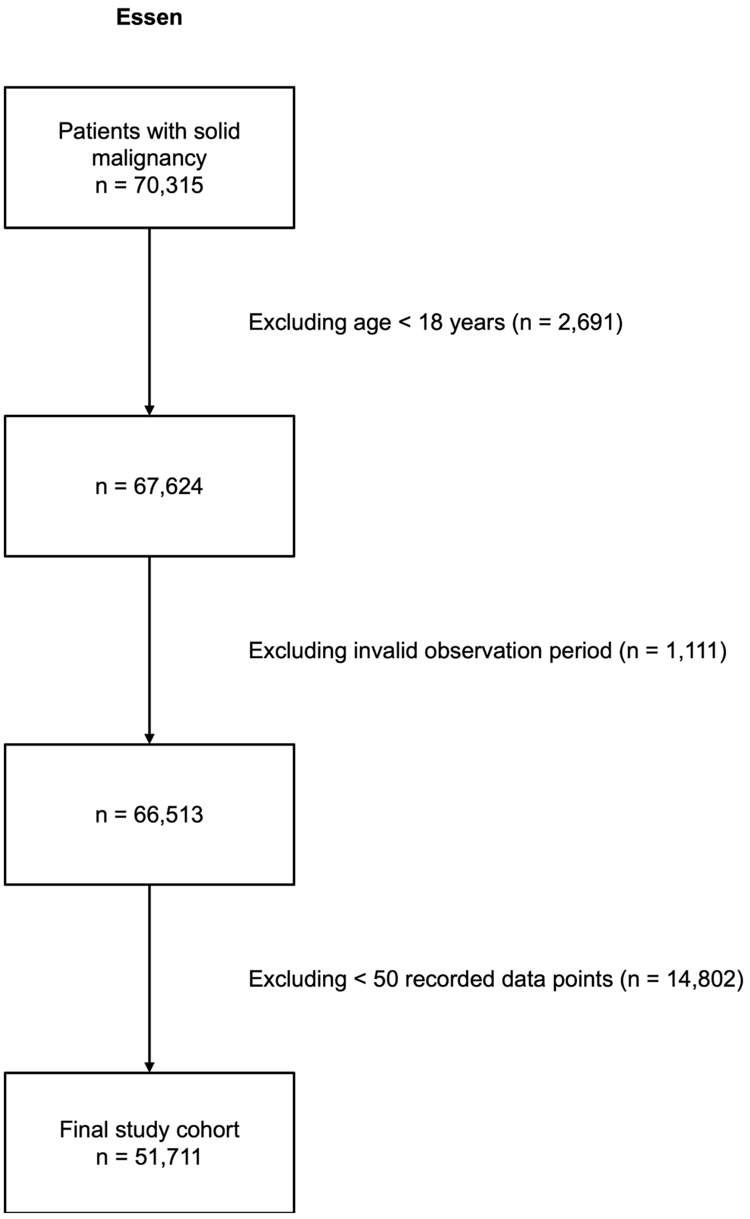


**Supplementary Figure 1:** Flow Chart showing data preprocessing.

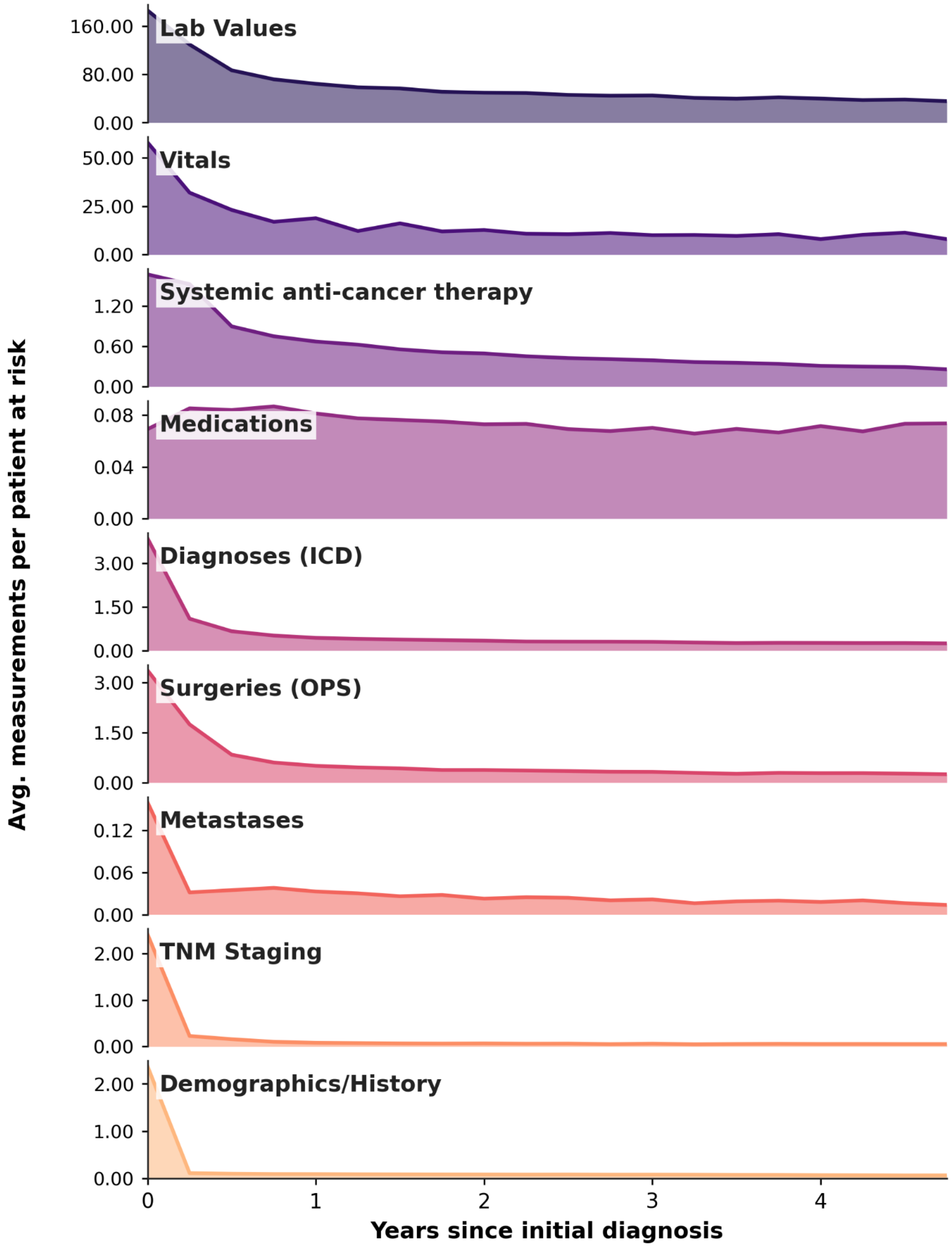


**Supplementary Figure 2: Data density by feature group over time.** Average measurements per patient at risk in 3-month bins over 5 years following initial diagnosis (51711 patients, Essen cohort). Counts in each bin were normalized by the number of patients at risk.

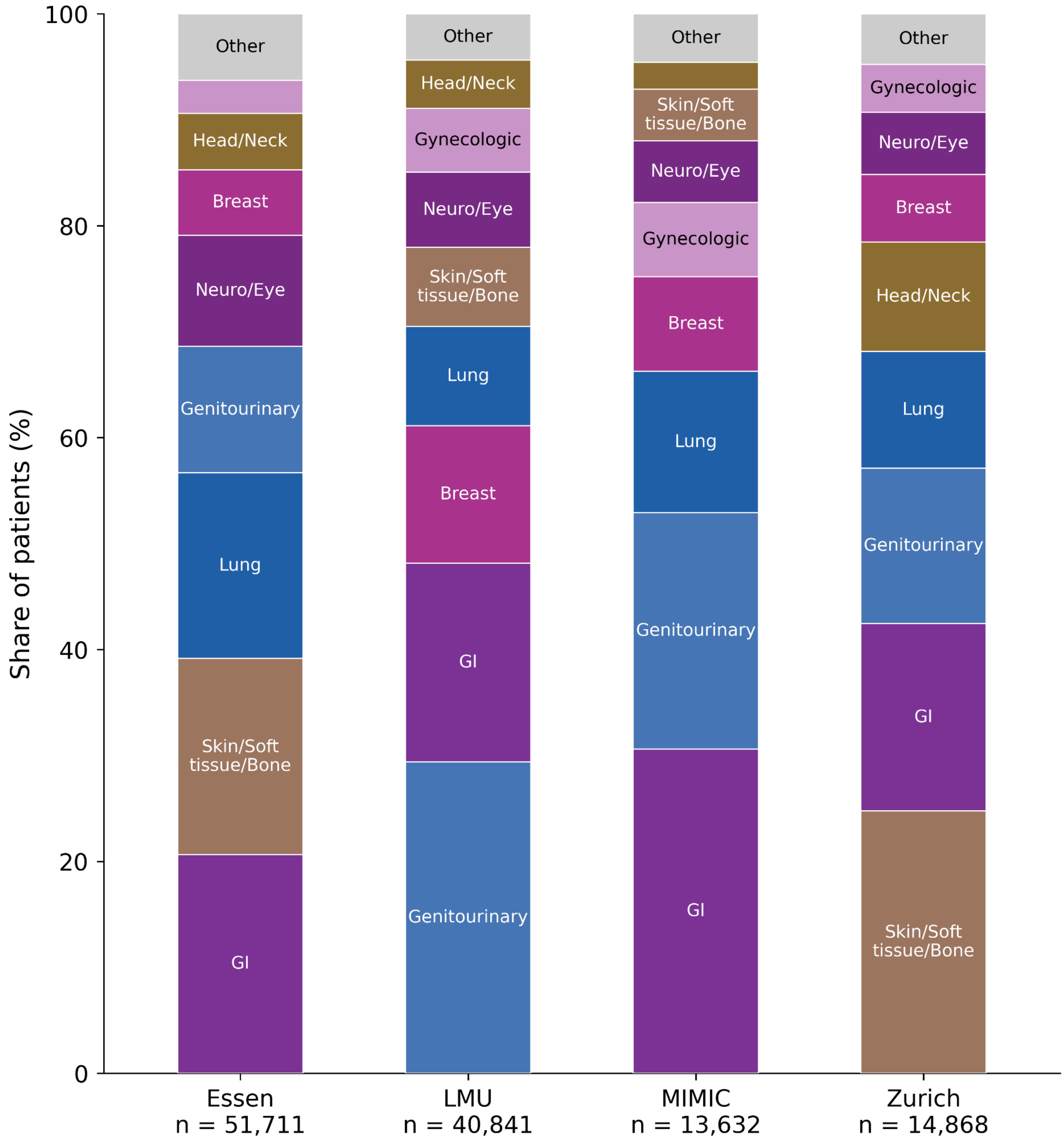


**Supplementary Figure 3: Distribution of primary tumor sites across the internal training cohort and external validation cohorts.** Stacked bars showing the share of patients per primary tumor site in the internal training cohort (Essen) and the three external validation cohorts (LMU, MIMIC-IV, Zurich). Primary tumor sites were derived from ICD-10 codes recorded at the time of initial diagnosis and grouped into anatomical categories. The eight most prevalent categories across cohorts (ranked by mean share) are shown individually; remaining categories are aggregated into "Other". Within each bar, categories are stacked from most to least prevalent (bottom to top). Patient numbers per cohort are indicated below each bar.

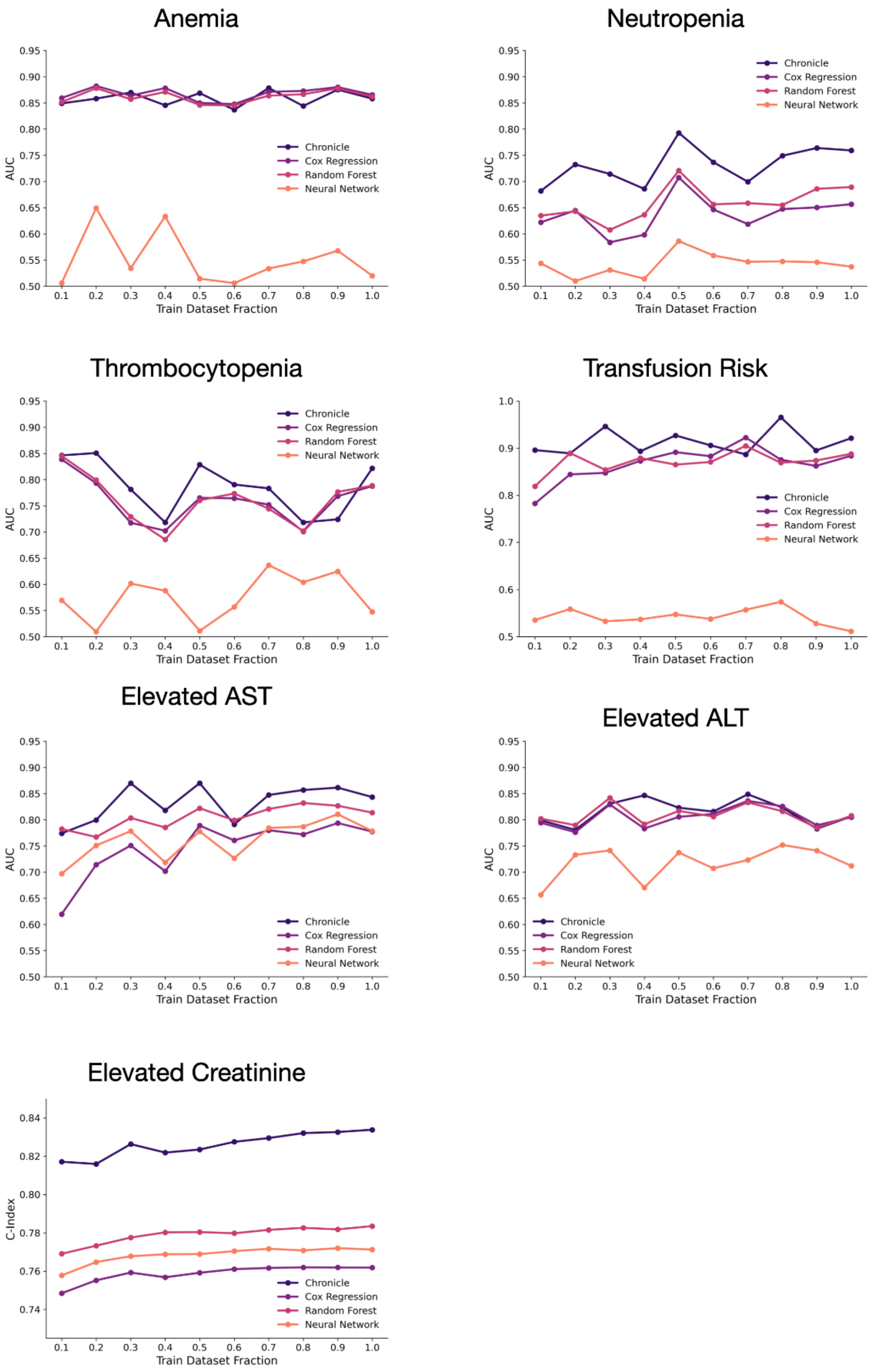


**Supplementary Figure 4:** Scaling study assessing model performance according to training dataset size. Models were trained on increasing fractions of the dataset (10% to 100%).

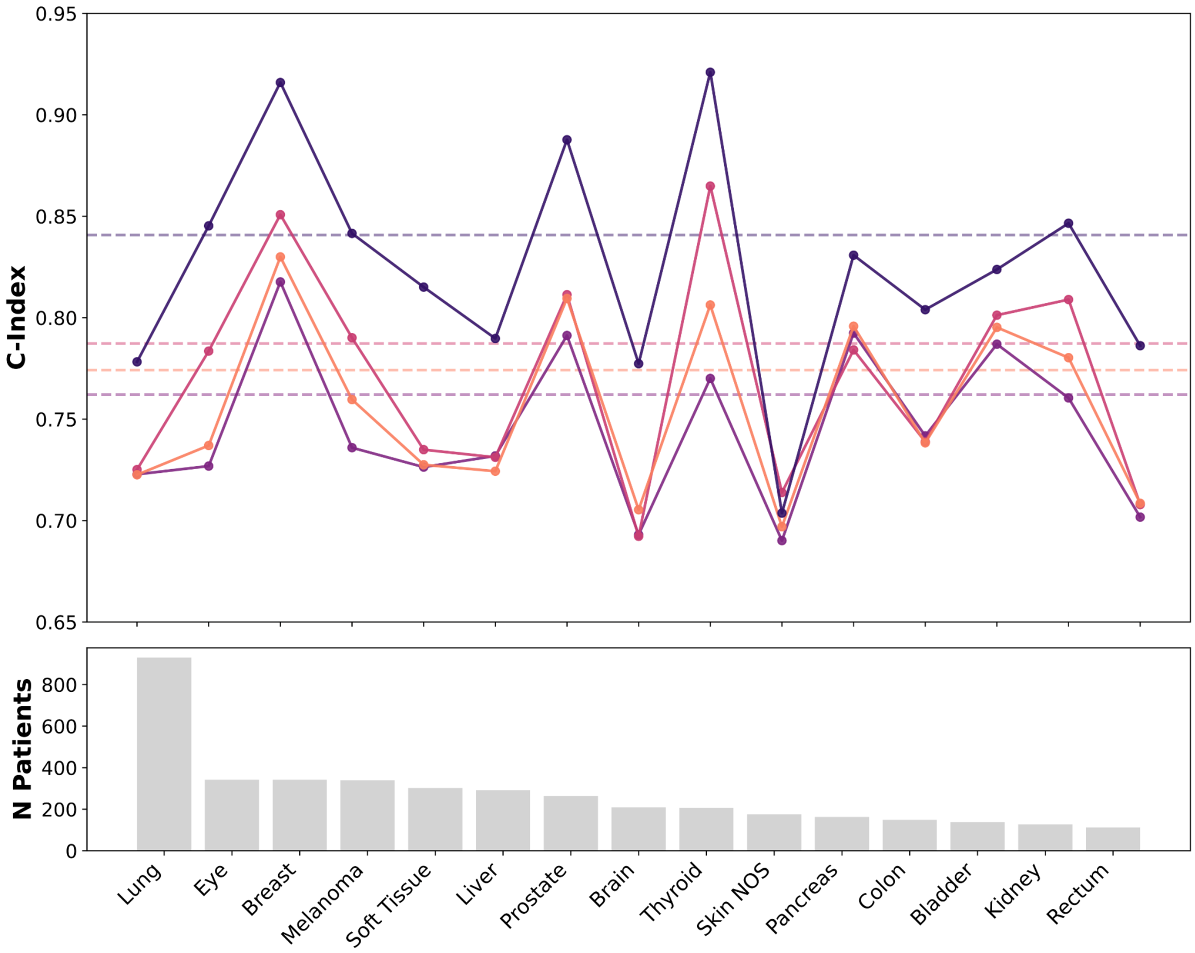


**Supplementary Figure 5:** Comparison of C-indices for overall survival prediction across different tumor entities. Chronicle is compared against conventional cross-sectional approaches (Cox Proportional Hazards regression, Neural Network, Random Survival Forest). Each point represents performance within a specific cancer type, with dashed lines indicating the mean performance across all entities. The bar plot below shows the number of patients per tumor group.

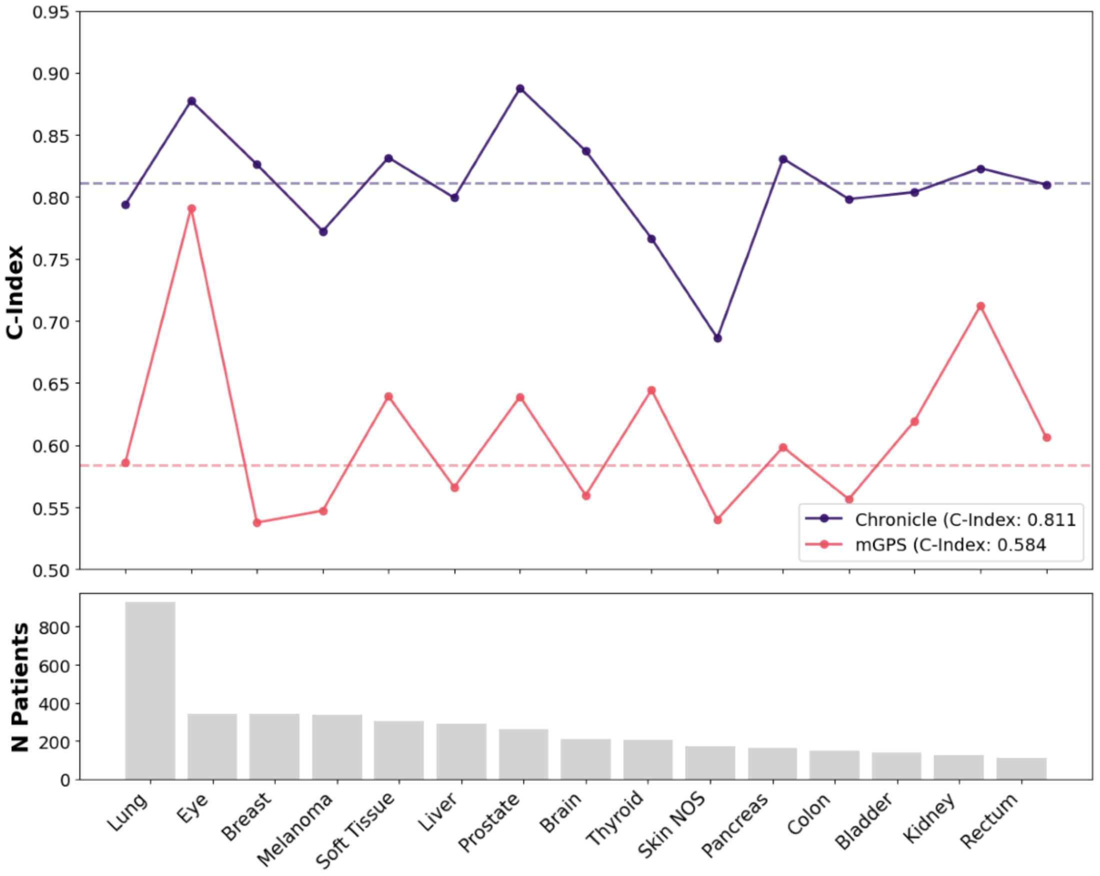


**Supplementary Figure 6:** Comparison of C-indices for overall survival prediction across tumor entities. Chronicle is compared with mGPS score. Points represent performance per cancer type; dashed lines indicate mean performance across entities. The bar plot displays patient numbers per tumor group.

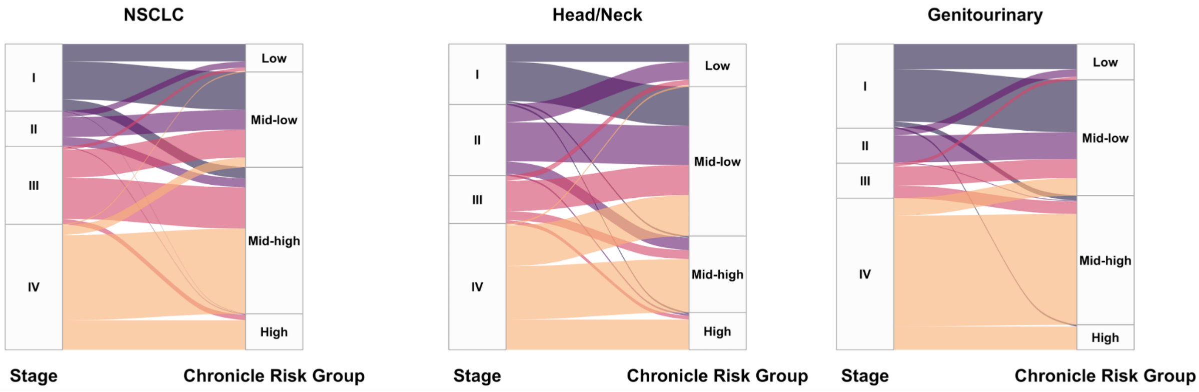


**Supplementary Figure 7: Stage-to-risk-group reclassification across cancer cohorts.** Alluvial diagrams showing the redistribution of patients from clinical stage (left, I-IV) to Chronicle model-predicted risk group (right: Low, Mid-low, Mid-high, High) for (a) NSCLC, (b) Head/Neck, and (c) Genitourinary cohorts. Node height is proportional to the number of patients in each stage or risk category; ribbon width represents the number of patients following that stage-to-risk-group transition, and ribbons are colored by stage of origin.

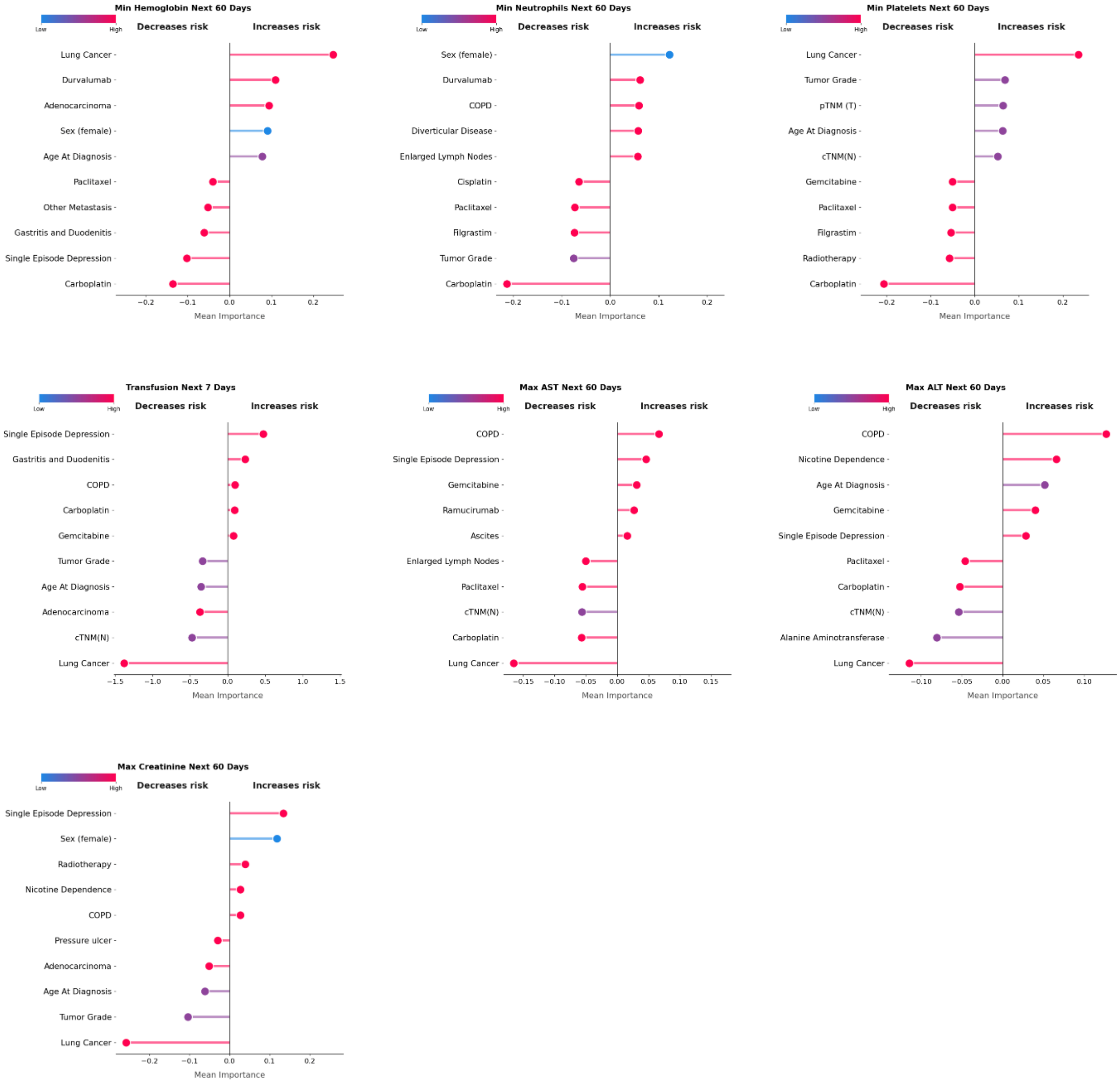


**Supplementary Figure 8:** A dot plot depicting mean LRP importance (x-axis) across the full cohort for individual clinical variables (y-axis) across the seven prediction tasks (60-day nadir hemoglobin; neutrophils; platelets; transfusion risk; elevated aspartate aminotransferase, elevated alanine aminotransferase and elevated creatinine over 60 days). Points are color-coded by feature value (blue: low, red: high). Features are ordered by their contribution to decreasing (left) or increasing (right) predicted risk.

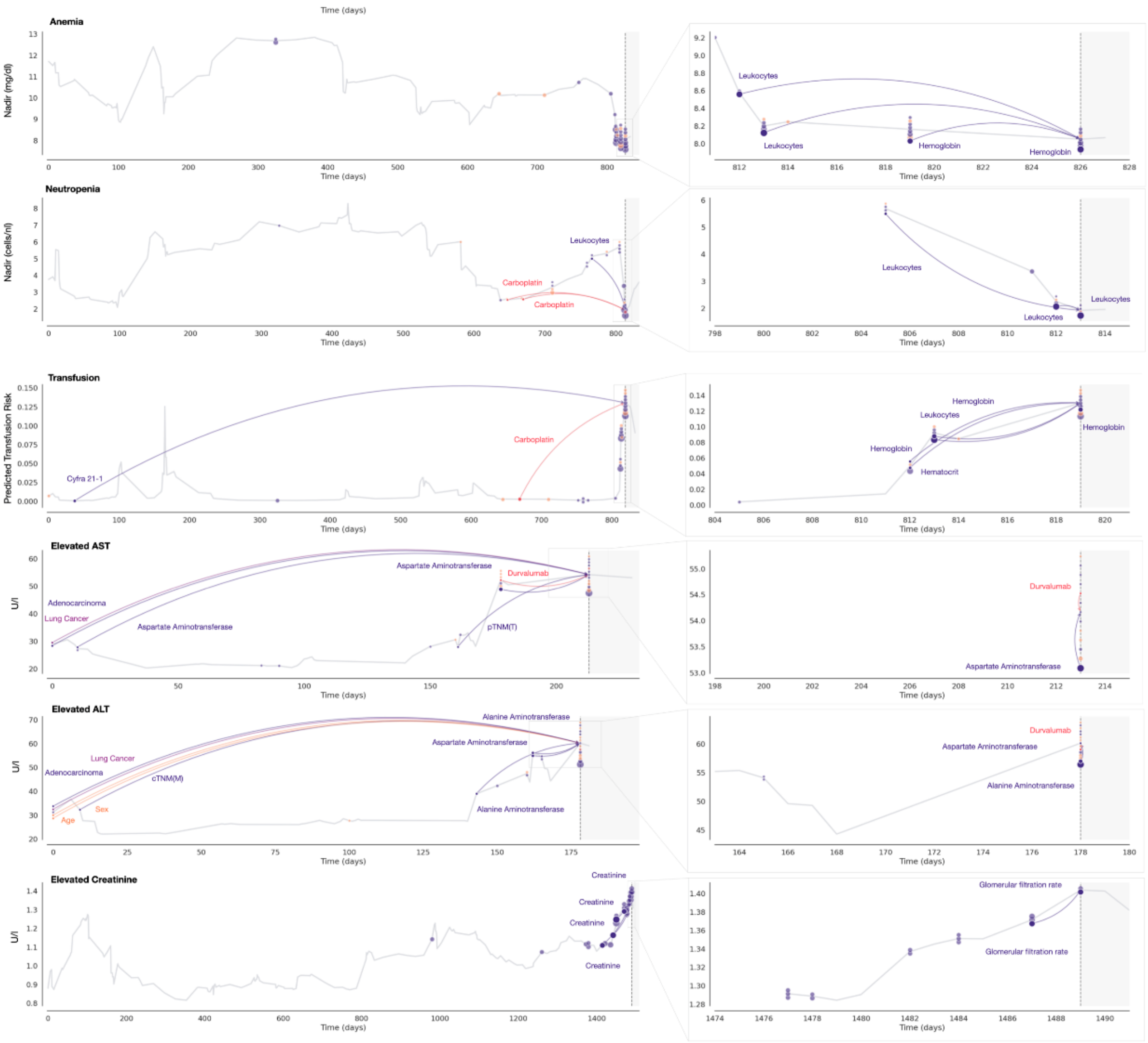


**Supplementary Figure 9:** Longitudinal LRP-based explanations for 60-day nadir of hemoglobin and neutrophils, transfusion risk, 60-day elevated AST, ALT and creatinine prediction. The left panel shows the prediction trajectory until the explained timepoint (dashed vertical line), with arrows highlighting the strongest contributing patient characteristics. The right panel provides a zoomed view of the preceding period, emphasizing short-term temporal relevance. Variables are grouped into Observations, Diagnoses, Interventions, and Other.

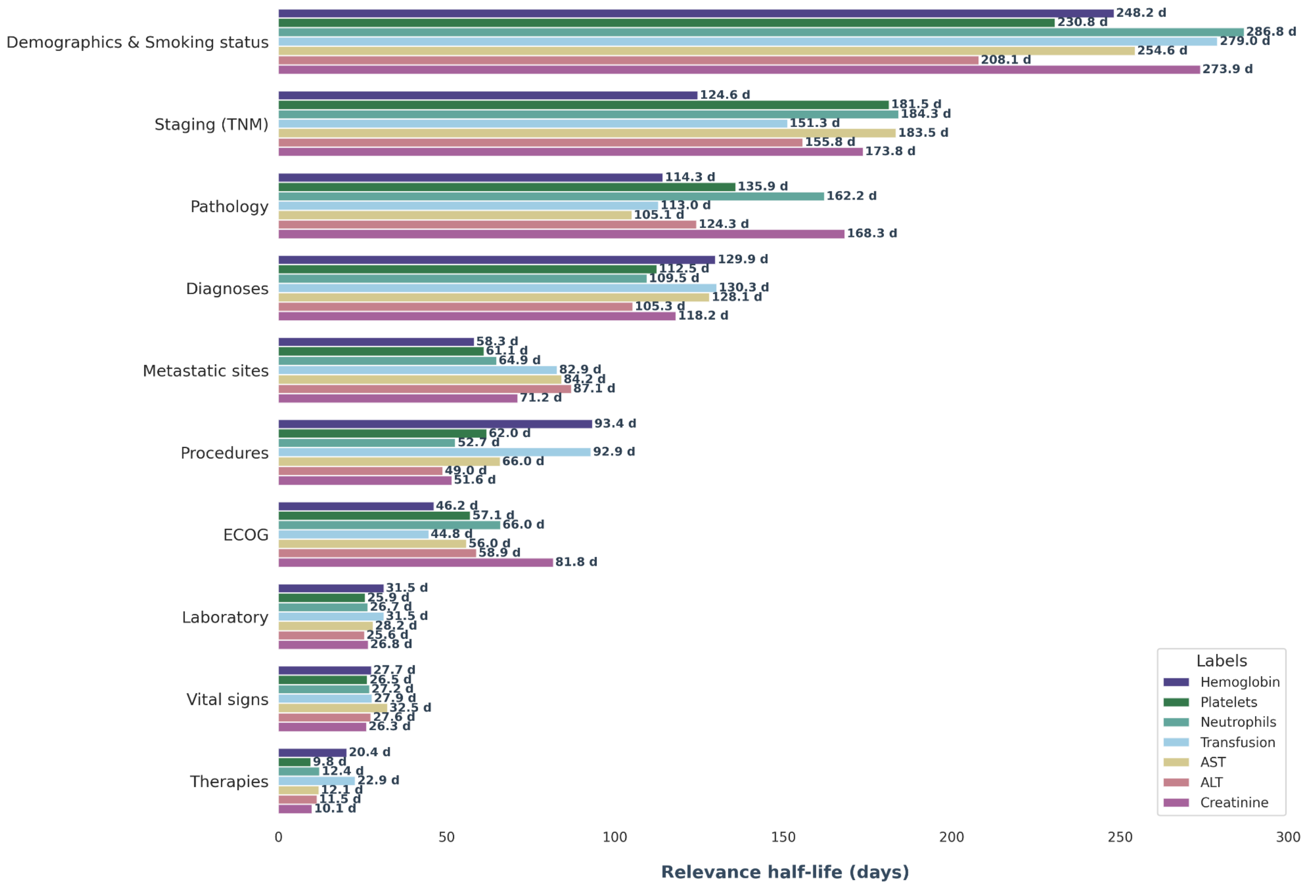


**Supplementary Figure 10:** Analysis of variable relevance decay, showing the temporal stability of information across different feature groups and labels for a subset of 1000 patients per label. The x-axis denotes the mean number of days until the relevance of a measurement decreases by 50%.

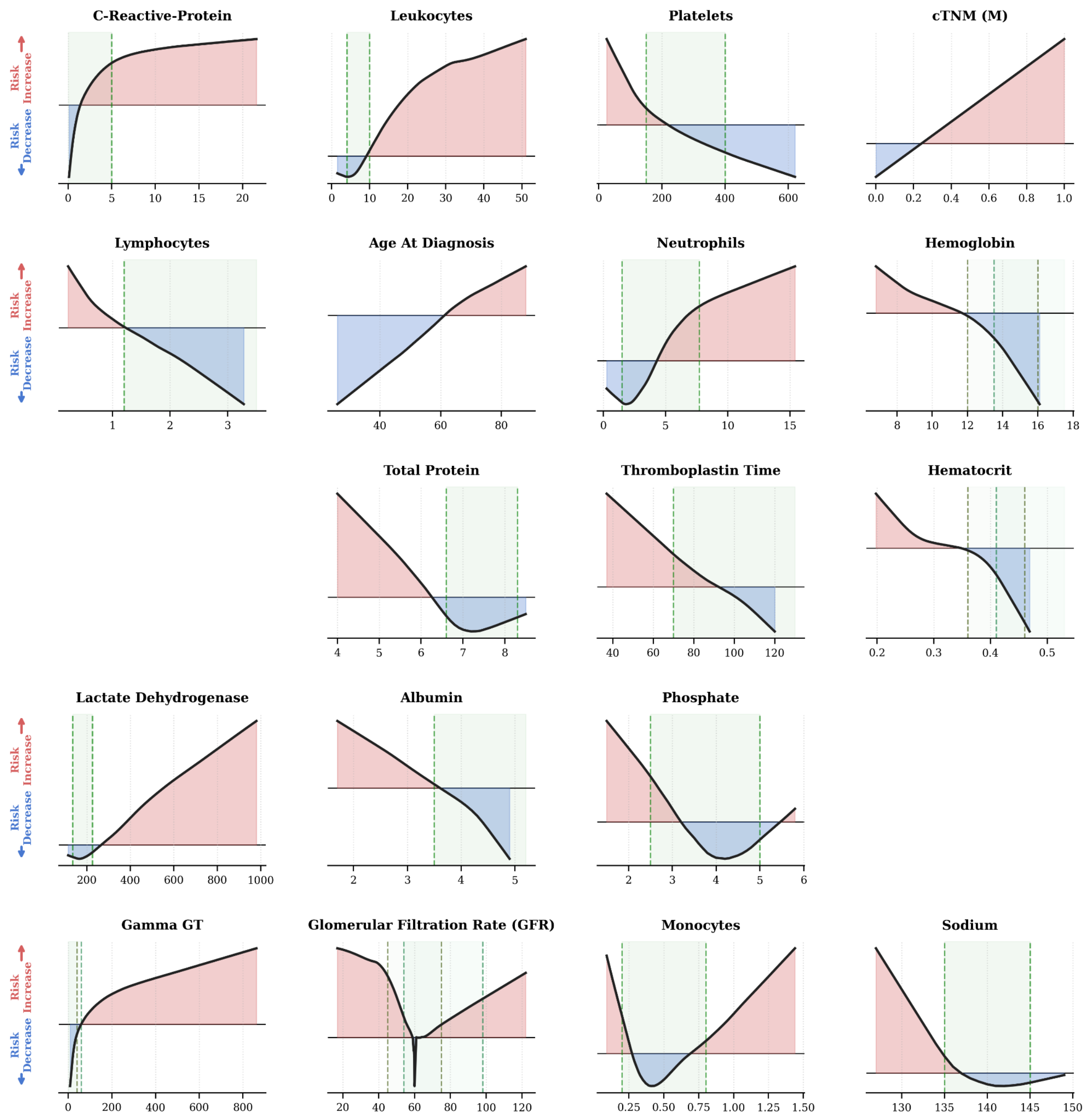


**Supplementary Figure 11:** Relationship between individual variable values (x-axis) and their corresponding mean LRP-based risk contributions (y-axis) for the independent Munich cohort. Predictions were generated in a zero-shot setting, the model trained on the Essen cohort was applied directly to the previously unseen Munich cohort, without any fine-tuning on Munich data. Positive values (red) indicate a higher than average predicted risk, while negative values (blue) indicate a lower than average predicted risk. The physiological reference ranges for each laboratory parameter are highlighted by green dashed vertical lines and shaded areas; when sex-specific differences exist, the reference ranges are shown separately for female (light green) and male (dark green) patients.
